# Novel discoveries and enhanced genomic prediction from modelling genetic risk of cancer age-at-onset

**DOI:** 10.1101/2022.03.25.22272955

**Authors:** Ekaterina S. Maksimova, Sven E. Ojavee, Kristi Läll, Marie C. Sadler, Reedik Mägi, Zoltan Kutalik, Matthew R. Robinson

## Abstract

Genome-wide association studies seek to attribute disease risk to DNA regions and facilitate subject-specific prediction and patient stratification. For later-life diseases, inference from case-control studies is hampered by the uncertainty that control group subjects might later be diagnosed. Time-to-event analysis treats controls as right-censored, making no additional assumptions about future disease occurrence and represents a more sound conceptual alternative for more accurate inference. Here, using data on 11 common cancers from the UK and Estonian Biobank studies, we provide empirical evidence that discovery and genomic prediction are greatly improved by analysing age-at-diagnosis, compared to a case-control model of association. We replicate previous findings from large-scale case-control studies and find an additional 7 previously unreported independent genomic regions, out of which 3 replicated in independent data. Our novel discoveries provide new insights into underlying cancer pathways, and our model yields a better understanding of the polygenicity and genetic architecture of the 11 tumours. We find that heritable germline genetic variation plays a vital role in cancer occurrence, with risk attributable to many thousands of underlying genomic regions. Finally, we show that Bayesian modelling strategies utilising time-to-event data increase prediction accuracy by an average of 20% compared to a recent summary statistic approach (LDpred-funct). As sample sizes increase, incorporating time-to-event data should be commonplace, improving case-control studies by using richer information about the disease process.

## Introduction

Cancer has broad medical importance and a high global health burden, with 19.3 million new cancer cases and almost 10 million cancer deaths occurring in 2020 [1]. Genome-wide association studies (GWAS) aim to attribute risk to regions of the DNA [2] and facilitate polygenic risk score (PRS) calculation [3] to predict subject-specific risk, which may then enable targeted and improved healthcare [4–6]. There is currently evidence for only 450 genomic regions associated with increased risk of 18 common cancers [2], despite recent results showing significant non-zero heritability across a range of cancer occurrences [7]. Current PRS calculated from these findings stratify risk for several cancers, including breast, colon, and prostate cancer, but often add negligible additional predictive information compared to existing non-genomic predictors [8]. Increasing sample size yields increased statistical power for discovery, with extensive recent case-control studies for breast cancer [9], prostate cancer [10, 11], ovarian cancer [12], or testicular cancer [13] showing improved results, but this remains a challenging endeavour. Biobanks provide an additional resource, essential for modern-day medical genetics; however, individuals within these studies have not all reached old age and the number of cancer cases is not high, with recent studies combining biobank cohorts for 18 cancer types [7] to limited effect.

Increased statistical power can also stem from tailored modelling choices, and one factor behind limited predictive performance could be the choice of the genome-wide analysis method. Although most association studies use methods that account for the impact of other genetic regions (fastGWA [14], GMRM [15], BoltLMM [16], REGENIE [17]), it is sometimes still preferred to resort to the basic association testing. In addition, most genome-wide analyses have been performed using a case-control phenotype rather than utilising the cancer diagnosis age as a phenotype, and there is some evidence that analysing data using time-to-event informed methods can have more power for detecting associations [18–21].

Here, we provide empirical evidence using data on 11 common tumours from the UK and Estonian Biobank studies that GWAS discovery and genomic prediction are greatly improved by analysing age-at-diagnosis, compared to a case-control model of association. We extend our recently presented BayesW approach [20], a Bayesian modelling framework that enables joint effect size estimation for time-to-event data, to provide marginal leave-one-chromosome-out mixed-linear age-at-onset adjusted association estimates, in contrast to using Cox mixed model [22] or age-at-onset informed genomic reconstruction of the phenotype [21]. We focus on a re-analysis in the UK Biobank data alone, and we replicate previous findings from large-scale case-control GWAS and find an additional 7 previously unreported independent genomic regions, out of which 3 replicated in independent data. Our novel discoveries provide new insights into underlying cancer pathways, and our model yields a better understanding of the polygenicity and genetic architecture of the 11 tumours. We find substantial SNP-heritability, implying that heritable germline genetic variation plays a vital role in cancer occurrence, with risk attributable to many thousands of underlying genomic regions. Finally, we show that Bayesian modelling strategies that utilise time-to-event data give increased prediction accuracy for all analysed tumours and suggest clinically relevant discrimination rules within the Estonian Biobank study. We argue that it is possible to use existing data more thoughtfully and that a re-analysis of case-control study data exploiting age-at-onset information would lead to novel discoveries and enhanced genomic prediction within our framework.

## Results

We analysed data from the UK Biobank for the timing and occurrence of diagnosis of 11 different tumours using 458,747 individuals of European ancestry and a very weakly LD pruned set of 2,174,071 SNP markers (see Methods and descriptive statistics in Supplementary table S1). We applied the recently developed GMRM-BayesW and GMRM-BayesRR-RC approaches to the age-at-onset and case-control data respectively in order to obtain joint SNP marker effect estimates for both models. We used these joint effect size estimates to describe the genetic architecture of the 11 cancers and to predict cancer occurrence within the Estonian Biobank data. Additionally, we adapted REGENIE’s mixed-linear association model by providing leaving one chromosome out (LOCO) predictors from the joint GMRM-BayesW and GMRM-BayesRR-RC analyses in step 2 of REGENIE (see Methods). This procedure allowed us to obtain marginal summary statistics, which we used to identify novel associations.

### Genetic architecture of 11 cancers

First, we used the power of joint Bayesian modelling to describe the genetic architecture of 11 cancers, whilst accounting for the MAF and LD structure of the genetic markers. We find that all traits are highly polygenic, with most of the 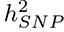 attributed to SNPs that contribute an average of 0.1% and 0.01% of the group genetic variance for joint BayesRR-RC and BayesW models, respectively (Figure 1a). We find some differences across cancers, notably melanoma (10%), basal cell carcinoma (13%), breast (8.0%), cervical (5.1%), and prostate (5.4%) cancers; and for age-at-diagnosis of non-Hodgkin’s lymphoma (7.0%), bladder (6.0%) and ovarian (5.3%) cancers where at least 5% of the 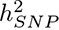 can be attributable to a small number of large effects (mixture 10^−2^) (Figure 1a). In general, the analysis of time-to-event phenotypes results in more of the genetic variance assigned to the smallest mixture component (Figure 1a). The result is in line with the number of LD-independent regions required to explain a proportion of the SNP heritability, where time-to-event analysis results in a more polygenic architecture compared to the case-control analysis (Figure 1b). Each curve reaches a plateau with 80-90% of the genetic variance attributable to a small number of genomic regions, and the remaining 10-20% attributable to 10,000 to 20,000 LD-independent regions. The number of remaining regions required to capture all of the association signals varies greatly across cancers, from 13,600 for ovarian and testicular cancer to 22,500 for basal cell carcinoma (Figure 1b). Additionally, we find that the 11 cancers differ in how rare and common variants contribute to the SNP heritability (Figure 1c). We further observe that genetic variance often positively correlates with variants’ MAF structure. For example, the largest proportion of genetic variance is consistently attributable to common variants in the fourth MAF quartile for both time-to-event (TTE) or case-control (CC) models on basal cell carcinoma (TTE 66%, CC 68%), melanoma (TTE 32%, CC 36%), breast (TTE 44%, CC 70%), colon (TTE 43%, CC 36%), and prostate cancers (TTE 40%, CC 57%) (Figure 1c). In contrast, testicular cancer, non-Hodgkin’s lymphoma and ovarian cancer have 61%, 54%, 63% of the genetic variance explained by the rarest effects in the first MAF quartile according to the case-control model. Thus, our MAF-LD stratified 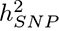 estimation approach suggests: (i) strong differences in the underlying genetic architecture across these 11 cancers, (ii) that only a limited number of genomic regions are required to capture most of the risk for all cancers, and (iii) that mapping further associations will be extremely difficult as a small amount of variance is attributable to a large number of independent regions of the DNA.

**Figure 1.**
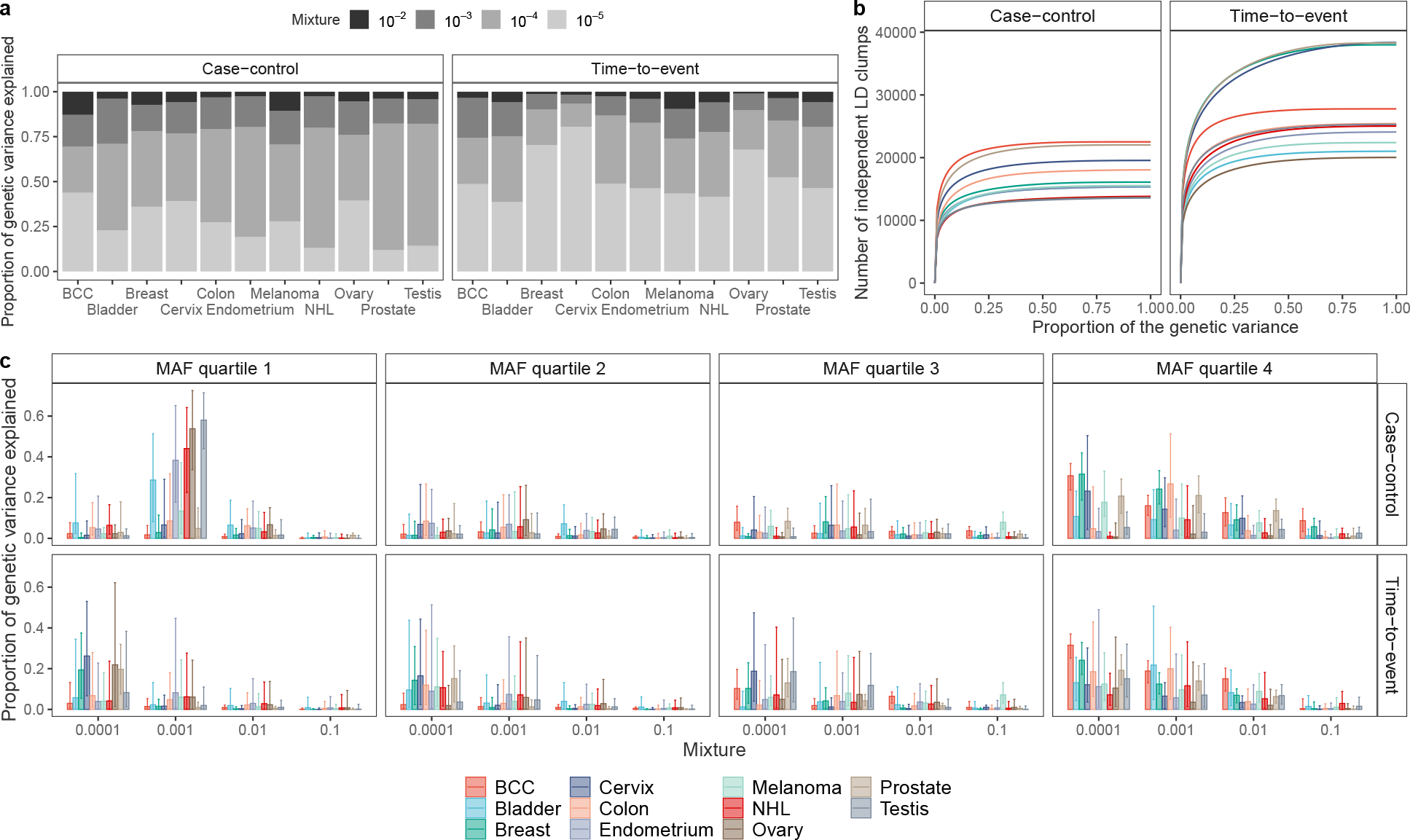
Genetic architecture and polygenicity of 11 cancers. (a) Mean proportion of genetic variance explained by each of the mixtures components using either case-control or age-at-onset phenotype. We find evidence that age-at-onset is highly polygenic with most of the genetic variance attributable to SNPs contributed by markers in the 10^−4^ mixture group, while the majority of the case-control phenotype genetic variance is explained by the markers from the 10^−3^ mixture group. (b) Number of LD-independent regions (see Methods) needed to explain total genetic variance. The contributions of LD-independent regions were sorted ascendingly such that the smallest contributing regions were added first. (c) Median proportion of genetic variance explained by each mixture class and MAF quartile combination, with 95% CI. For both case-control and age-at-onset models, most of the genetic variance is attributable to the small effect common variants (MAF quartile 4), however rare variants from the first MAF quartile contribute significantly to the variance for bladder, endometrial, ovarian, testicular cancers, non-Hodgkin’s lymphoma for the BayesR model. BCC indicates basal cell carcinoma.

We then aimed at estimating the genetic heritability of the 11 cancers using LD score regression [23] on the marginal associations. When correcting for the discrete nature of the trait (see Methods), the liability scale heritability estimates were closer to family-based estimates than array-based assays, especially for basal cell carcinoma, melanoma, breast, prostate, and testicular cancers (Table 2), highlighting that heritable genetic variation is a leading risk factor for underlying risk of cancer. The pattern holds even if we use an approach tailored for estimating liability scale heritability for rare traits [24] resulting in slightly more conservative estimates (Table S3). Interestingly, we find that the GMRM-BayesW analysis leads to heritability estimates that are nominally identical to the GMRM-BayesRR-RC estimates, suggesting an equivalent description of total genetic variance when including the timing information in the analysis. The joint Bayesian models for occurrence also enable SNP heritability estimation and comparative inference across cancers of the underlying distribution of genetic effects. The liability scale heritability estimates from the joint Bayesian model are less conservative and are similar to the LD score regression analysis estimates for more prevalent cancers. However, more remarkable differences between the estimates and wider credibility intervals occur for the less prevalent cancers, supporting suggestions [24] that rare traits require extra care as they could be subject to ascertainment bias, sampling bias, and their effective sample size is low. We further used cross-trait LD score regression on the GMRM-BayesW or GMRM-BayesRR-RC adjusted marginal associations to estimate the genetic correlation between the traits (see Methods). There is a sizable genetic correlation between melanoma and basal cell carcinoma (GMRM-BayesW adjusted estimate 0.44, 95%CI (0.32, 0.57)), as well as multiple significant genetic correlations between basal cell carcinoma, cervical cancer and other phenotypes (Table S4). Altogether, Bayesian modelling provides heritability estimates that are close to family-based assays, especially for prevalent cancers, and allows to recover genetic variation underlying the risk of cancer both with case-control and time-to-event data.

The joint Bayesian models for occurrence also enable SNP heritability estimation and comparative inference across cancers of the underlying distribution of genetic effects. The liability scale heritability estimates from the joint Bayesian model are similar to the LD score regression analysis estimates for more prevalent cancers. However, more remarkable differences between the estimates and wider credibility intervals occur for the less prevalent cancers, supporting suggestions [24] that rare traits require extra care as they could be subject to ascertainment bias, sampling bias, and their effective sample size is low. We further used cross-trait LD score regression on the BayesW or BayesRR-RC adjusted marginal associations to estimate the genetic correlation between the traits (see Methods). There is a sizable genetic correlation between melanoma and basal cell carcinoma (BayesW adjusted estimate 0.51, 95%CI (0.34, 0.68)), and we replicate [7] a previous result of negative genetic correlation between endometrial and testicular cancer (BayesW adjusted estimate -0.38, 95%CI (-0.68, -0.07)) (Table S4). Interestingly, BayesW-based genetic correlations have a narrower confidence interval than BayesRR-RC based genetic correlation estimates for each significant cancer trait pair.

### Genomic prediction of 11 cancers

Next, we used joint GMRM-BayesW SNP marker estimates to predict cancer occurrence within the Estonian Biobank data (Figure 2, see Methods). We compared our results to those obtained by a baseline joint GMRM-BayesRR-RC model and also to LDpred-funct approach [25] that uses summary statistics from fastGWA method [14] for the same variants using precisely the same individuals as for the Bayesian models (see Methods). Furthermore, we provide a comparison of applying Bayesian models on either self-reported or medical record data (see Methods), showing that medical record data models outperform models using self-reported data. Thus, we resorted to using only medical record data (Figure S4), illustrating the importance of high data quality and accurate measurement to facilitate phenotype linking across studies.

**Figure 2.**
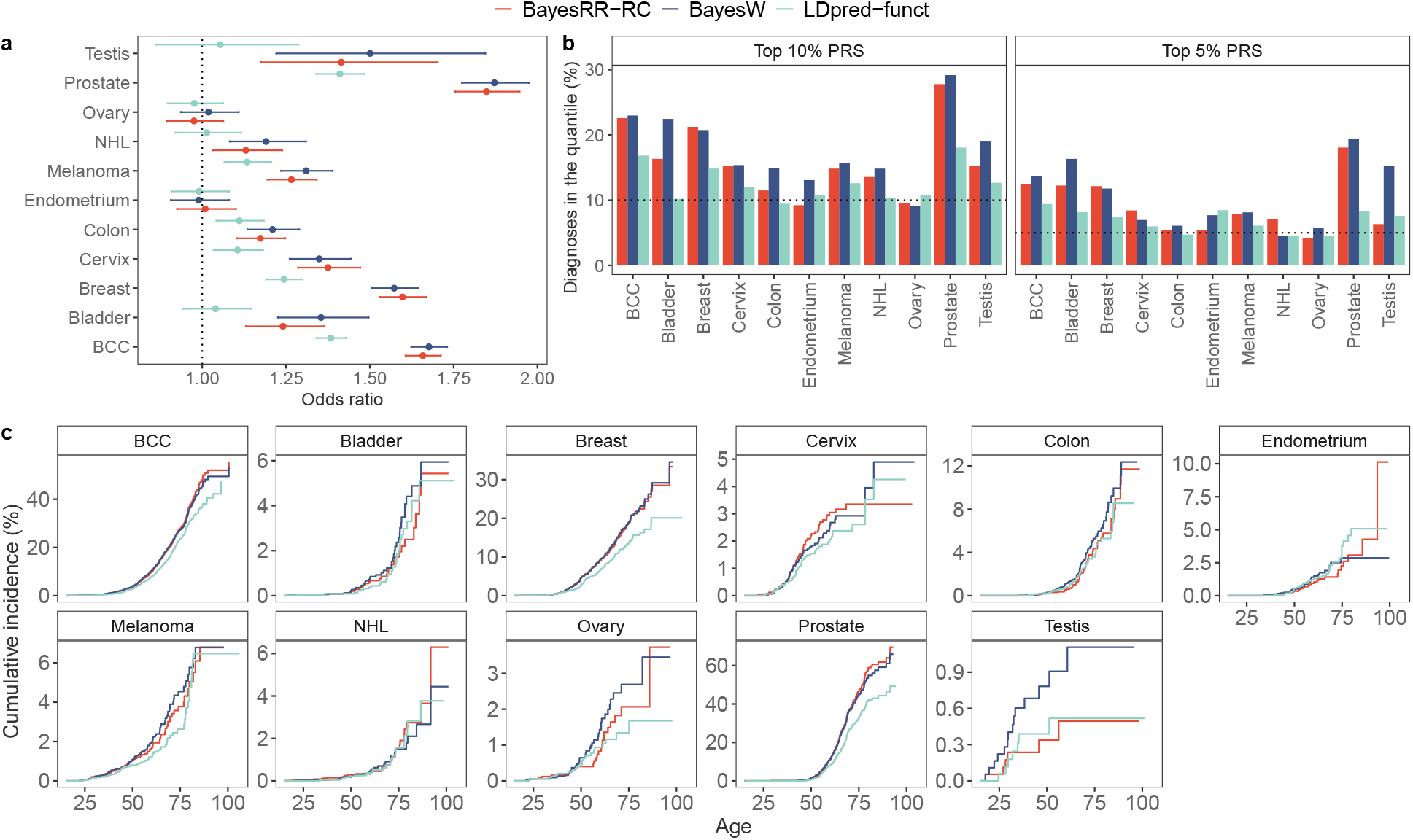
Predictive validation of different polygenic risk scores (PRS) in the Estonian Biobank data. (a) Odds ratio for diagnosis of a tumour given one standard deviation increase in PRS, with 95% confidence intervals. (b) Percent of individuals diagnosed with cancer before age 50 having a top 10% or top 5% highest PRS; (c) cumulative incidence curves adjusted for competing risk for individuals with the top 5% highest PRS. The number of Estonian Biobank individuals used in the validation was *N* =195,432. BayesRR-RC and BayesW estimates were obtained by running the corresponding models on UK Biobank using either case-control or age-at-onset data. The LDpred-funct used the summary statistics that were calculated using the same UK Biobank individuals and variants as for BayesRR-RC or BayesW using fastGWA method; then the summary statistics were used in the LDpred-funct method [25] (see Methods). BayesRR-RC and BayesW tend to have more accurate predictions than the summary statistic approach. For all cancers except breast, cervical, endometrial and ovarian cancer, BayesW predictor gives a nominally higher odds ratios compared to BayesRR-RC predictor.

We find that joint Bayesian models for individual-level data, especially those utilising age-at-onset information, yield substantially improved genomic prediction for cancer occurrence, and the benefit is amplified as case count increases. Except for a few cancers, we find that conducting the analysis using an age-at-onset phenotype (BayesW) yields a nominally higher odds ratio (of having one standard deviation higher PRS) than a case-control phenotype (BayesRR-RC) for basal cell carcinoma (BayesW: 1.68, BayesRR-RC: 1.66), bladder cancer (BayesW: 1.35, BayesRR-RC: 1.24), colon cancer (BayesW: 1.21, BayesRR-RC: 1.17), melanoma (BayesW: 1.31, BayesRR-RC: 1.27), non-Hodgkin’s lymphoma (BayesW: 1.19, BayesRR-RC: 1.13), prostate cancer (BayesW: 1.87, BayesRR-RC: 1.85), and testicular cancer (BayesW: 1.50, BayesRR-RC: 1.41) (Figure 2a). A similar trend can be observed when using the C-statistic or hazards ratio for comparison (Figure S3, see Methods). The Bayesian approaches generally have better predictive accuracy than the summary statistic approach using LDpred-funct, with an average of 20% increase in odds ratio. LDpred-funct yielded at least nominally smaller odds ratios (of standard deviation PRS difference) for testicular cancer (LDpred-funct: 1.05), prostate cancer (LDpred-funct: 1.41), breast cancer (LDpred-funct: 1.24) and non-Hodgkin’s lymphoma (LDpred-funct: 1.01) with differences odds ratios differing significantly between BayesW and LDpred-funct for bladder, prostate and breast cancers. For the cases of non-Hodgkin’s lymphoma, bladder cancer and testicular cancer, the LDpred-funct score failed to yield a significantly predictive score.

We observe that the highest 5% PRS quantile discriminates well for the disease occurrence (Figure 2c). Whereas the risk of developing prostate cancer by age 85 is estimated to be 11% (Table S6) among the top 5% highest PRS individuals, nearly 58% or 61% will develop prostate cancer according to the BayesRR-RC or BayesW model, respectively. In comparison, LDpred-funct PRS finds that 43% of the top 5% PRS develop prostate cancer. The top 5% polygenic risk score yields a useful discrimination rule for most other cancers as well, notably for breast cancer, for which 25% of the top 5% BayesW PRS gets diagnosed by age 85 (12% in the population, Table S6) and basal cell carcinoma for which 44% of the top 5% BayesRR-RC PRS gets diagnosed by age 85 (31% in the population, Table S6). The share of individuals getting a cancer diagnosis before age 50 is disproportionately higher among individuals with the top 5% or 10% of the PRS across many cancers and risk score types (Figure 2b). For example, the top 10% highest genetic risk according to the BayesW risk score account for 23% out of all basal cell carcinoma cases and 29% out of all prostate cancer cases, suggesting that the BayesW risk score discriminates well the early onset of prostate cancer or basal cell carcinoma. Our results suggest that the top 5% highest Bayesian polygenic risk scores could serve as a rule to detect individuals who should not only receive earlier communication about their risks, but it could also result in a cost-effective screening model for this subset of individuals.

### Novel and replicated associations

Then, in order to fine-map novel associations, we used marginal age-at-onset and case-control summary statistics that we obtained by adjusting the mixed-linear association model REGENIE to run with joint LOCO predictors from GMRM-BayesRR-RC and GMRM-BayesW instead of built-in ridge LOCO predictors [15,17,20] (see Methods). We find that this approach of adjusting the phenotypes with joint Bayesian predictors results in enhanced statistical power (Figures 3c, S5, S1, S2,). We observe particularly notable improvements in p-values for melanoma, bladder and testicular cancers.

**Figure 3.**
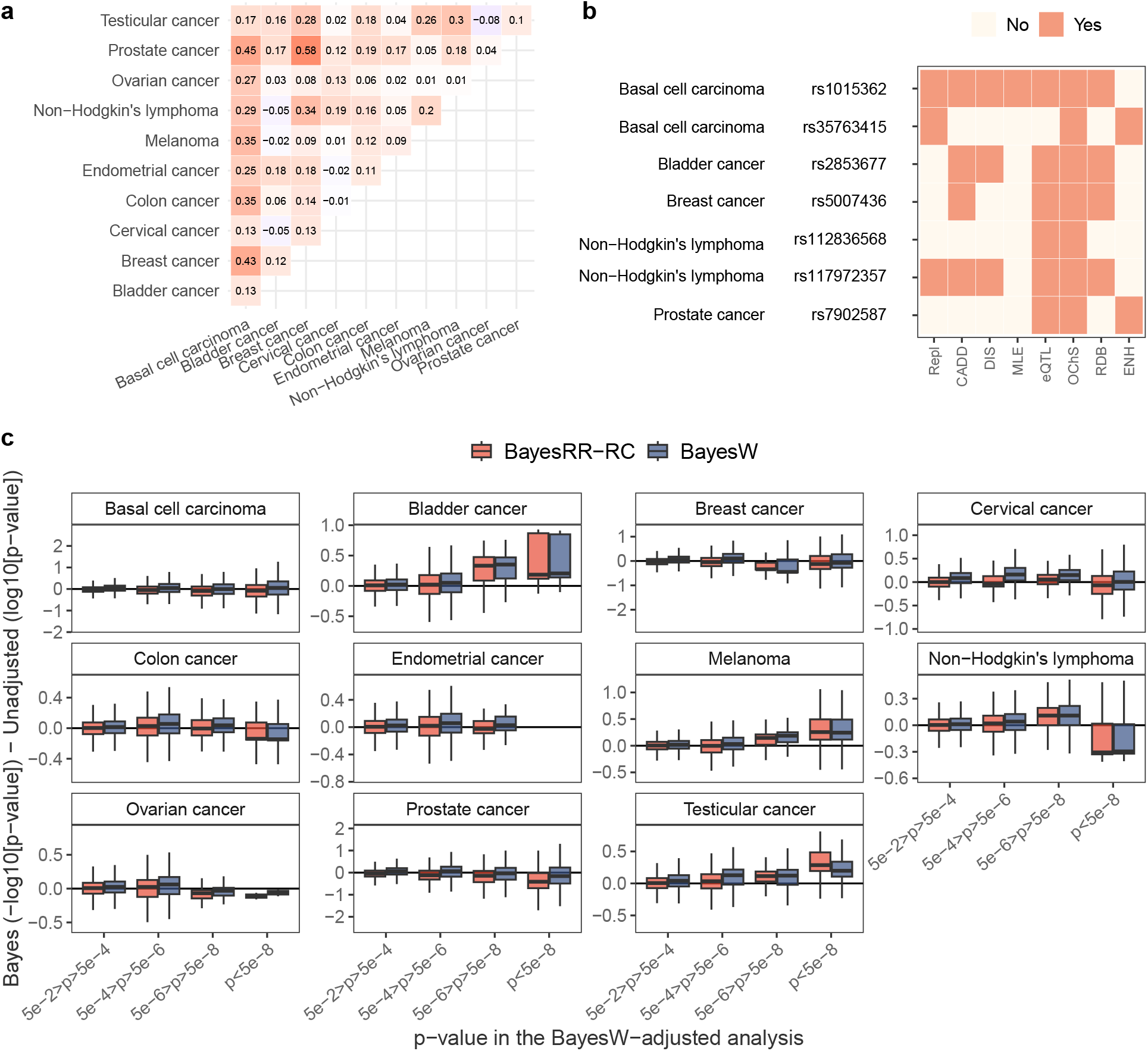
Properties of discoveries and changes in p-values. (a) Pearson correlation between key gene scores. Correlations were calculated using all key genes (including non-significant ones). (b) Properties of novel replicated genetic regions. Repl - the discovery was replicated in the Estonian Biobank, CADD - maximum CADD score of the region is equal or greater than 12.37, DIS - maximum DeepSEA disease impact score (DIS) of the genetic region is equal or greater than 2, MLE - maximum DeepSEA mean log e-value (MLE) of the region is equal or greater than 2, eQTL - an SNP from the the genetic region is an eQTL with p-value < 5 · 10^−8^, OChS - open/active chromatin state (minimum 15-core chromatin score of the lead SNP is less or equal than 7), RDB - minimum RegulomeDB category of the genetic region is 1 or 2, ENH - SNP is in enhancer region. (c) Differences between p-values from standard REGENIE analysis and BayesW- or BayesRR-RC-adjusted analyses. With the exception of two classes in non-Hodgkin’s lymphoma and ovarian cancer (*p <* 5 · 10^−8^), the Bayesian adjustments yield similar or slightly improved results compared to standard REGENIE with notable improvements seen in bladder cancer, cervical cancer (5 · 10^−8^ *< p <* 5 · 10^−4^), melanoma, non-Hodgkin’s lymphoma (5 · 10^−8^ *< p <* 5 · 10^−6^) and testicular cancer.

Applying GMRM-BayesW or GMRM-BayesRR-RC to the UK Biobank data, we replicate previously reported findings, with 261 previously identified significant independent trait-marker associations at *p <* 5·10^−8^ (Supplementary data). We also find an additional 7 independent previously unreported variants significant at *p <* 5 · 10^−8^, of which 3 replicate in independent data of the Estonian Biobank (Figure 3b, Table 1, see Methods). We observe that all 7 previously undiscovered variants had small but not genome-wide significant p-values in the unadjusted analysis with REGENIE (see Methods), and using GMRM-BayesRR-RC or GMRM-BayesW adjustment reduced their p-values below a genome-wide significance threshold (Table S5). We discover novel or replicate previous discoveries slightly better when we account for age-at-onset as compared to the case-control model, especially for traits that have higher case counts such as breast or prostate cancer (Figure S6, Table 1). Namely, only 3 out of 7 novel markers and 1 out of 3 replicated markers that were discovered with GMRM-BayesW adjustment were also discovered with GMRM-BayesRR-RC.

**Table 1.** Functional description of the novel and replicated discoveries from case-control (GMRM-BayesRR-RC) and age-at-onset (GMRM-BayesW) marginal analyses. We performed the marginal analysis by adjusting the mixed-linear association model REGENIE with GMRM-BayesW or GMRM-BayesRR-RC genetic LOCO predictors and identified 7 novel genetic loci, 3 of which were replicated in the Estonian Biobank (see Methods for the pipeline for filtering and replication of the novel loci). Importantly, two out of the three replicated loci were only discovered using the GMRM-BayesW adjusted model and one was discovered by using both GMRM-BayesW and GMRM-BayesRR-RC LOCO predictors. We calculated various parameters related to the potential functionality of novel genetic regions for each significant independent novel SNP and minimum/maximum/common values within the genetic regions (index SNPs and those in LD *r*^2^ *>* 0.6). The majority of SNPs from the 7 novel genomic regions could be linked to regulatory variation. Here, NHL - non-Hodgkin’s lymphoma, Heterochrom. - heterochromatin.

| SNP | rs1015362 | rs35763415 | rs2853677 | rs5007436 | rs112836568 | rs117972357 | rs7902587 |
| --- | --- | --- | --- | --- | --- | --- | --- |
| Replicated | False | True | False | False | False | True | True |
| Phenotype | Basal cell carcinoma | Basal cell carcinoma | Bladder cancer | Breast cancer | NHL | NHL | Prostate cancer |
| Model | bW,bR-RC | bW | bW | bW | bW,bR-RC | bW,bR-RC | bW |
| Eff/Oth | C/T | T/C | G/A | C/T | A/G | G/A | C/T |
| Chromosome | 20 | 17 | 5 | 18 | 18 | 14 | 10 |
| Position | 32738612 | 79622370 | 1287194 | 25437574 | 67608842 | 96043546 | 105694301 |
| Beta | -0.057 | 0.052 | -0.159 | 0.073 | 0.435 | 1.101 | 0.137 |
| SE | 0.010 | 0.009 | 0.029 | 0.013 | 0.074 | 0.171 | 0.025 |
| p-value | 4.255e-08 | 3.307e-08 | 4.885e-08 | 4.070e-08 | 4.222e-08 | 4.495e-08 | 4.482e-08 |
| Effect allele frequency | 0.278 | 0.590 | 0.577 | 0.749 | 0.028 | 0.003 | 0.095 |
| Index SNP CADD score | 0.261 | 1.09 | 1.28 | 0.886 | 6.518 | 8.4 | 0.032 |
| Max locus CADD score | 9.674 | 16.29 | 2.738 | 21.4 | 25.4 | 8.4 | 17.93 |
| Sei sequence class | AR | Polycomb/Heterochrom. | Polycomb | NANOG/FOXA1 | Low signal | CTCF-Cohesin | Erythroblast-like |
| Index SNP DeepSEA MLE | 0.446 | 0.425 | 0.631 | 0.243 | 0.301 | 0.664 | 0.394 |
| Max locus DeepSEA MLE | 1.207 | 1.264 | 0.786 | 1.042 | 1.412 | 0.851 | 2.553 |
| Index SNP DeepSEA DIS | 0.443 | 0.583 | 0.653 | 0.101 | 0.030 | 0.724 | 0.204 |
| Max locus DeepSEA DIS | 1.664 | 2.487 | 1.100 | 1.702 | 2.561 | 1.458 | 4.292 |
| ANNOVAR function | intergenic | intronic | intronic | intergenic | intronic | ncRNA intronic | intergenic |
| ANNOVAR functional gene | ASIP RPS2P1 | PDE6G | TERT | CDH2 RP11-739N10.1 | CD226 | RP11-1070N10.4 | OBFC1 SLK |
| Index SNP RDB |  | 5 | 5 | 7 | 5 | 3a | 5 |
| Min locus RDB | 5 | 2b | 5 | 2b | 2c | 3a | 2b |
| Min index SNP chromatin state | 5 | 2 | 4 | 5 | 3 | 2 | 5 |
| Min locus chromatin state | 1 | 1 | 4 | 1 | 1 | 1 | 1.0 |
| Common index SNP chromatin state | 15 | 5 | 15 | 15 | 15 | 14 | 15 |
| Common locus chromatin state | 15 | 4 | 14 | 15 | 15 | 5 | 5 |
| Chromatin interaction type | enhancer |  |  |  |  | enhancer |  |
| eQTL min p-value | 1.151e-88 | 3.272e-310 | 2.990e-08 | 7.441e-37 | 3.164e-33 |  | 1.858e-30 |

To map novel associations to functional annotations, we conducted a number of follow-up analyses (see Methods). 4 out of 7 novel lead SNPs are intronic (rs35763415 in PDE6G gene, rs2853677 in TERT, rs112836568 in CD226, and rs117972357 in ncRNA RP11-1070N10.4), while 3 other SNPs are intergenic (rs1015362, rs5007436, rs7902587). The majority of SNPs from the 7 novel genomic regions (lead SNPs and those in LD *r*^2^ *>* 0.6) could be linked to regulatory variation (Table 1). Namely, 6 out of 7 novel lead SNPs are expression quantitative trait loci (eQTLs) (maximum *p <* 2.9 · 10^−8^ from FUMA eQTL mapping, see Methods); 4 novel regions have SNPs that fall into RegulomeDB categories [26] that are likely to affect binding or are linked to expressions of a gene target (Figure 3b). Moreover, all 7 novel genomic regions are in open chromatin state in at least 1 of 127 tissue/cell types predicted by ChromHMM [27] (Figure 3b), while for one region (rs35763415), active chromatin state is the most common (Supplementary data). In addition, two novel lead SNPs are enhancers (rs1015362, rs117972357) (Figure 3b). Thus, most novel associations can be attributed to regulatory, intronic, and open chromatin functional regions.

We confirmed the regulatory effects of novel regions on a wide range of chromatin features using the DeepSEA Beluga and Sei models, deep-learning-based frameworks for systematical prediction of the chromatin effects of sequence alterations and of sequence regulatory activities [28, 29]. The Sei model predicted that 2 novel regions belong to the polycomb sequence class associated with repressive gene expression activity (rs35763415, rs2853677); rs1015362 maps to androgen receptor (AR) binding sequence which are reported to be involved in carcinogenesis and tumour growth in prostate cancer [30]; rs7902587 - to Erythroblast-like enhancer sequence class that is enriched in enhancer histone marks; enhancer SNP rs117972357 - to CTCF-cohesin sequence that regulates long-range genome interactions; and lastly, rs5007436 maps to NANOG / FOXA1 class that has cell type-specific transcription factor activity and has been implicated in tumour development [31]. The analysis with DeepSEA Beluga showed that one region (index SNP rs7902587, associated with prostate cancer) has maximum mean log e-value (MLE) > 2 (Figure 3b, Table 1), indicating a higher likelihood of regulatory effects than a reference distribution of 1000 Genomes variants. Moreover, this novel genetic region, as well as 2 other novel regions (rs35763415, rs112836568), has a maximum disease impact score (DIS) > 2 (Figure 3b, Table 1), highlighting likely disease-associated mutations. Additionally, we used the CADD tool that predicts deleterious, functional, and disease causal variants by integrating multiple annotations into one deleteriousness metric [32]. The average CADD score is 3.95, with 4 regions containing SNPs with max CADD score > 12.37 (Figure 3b, Table 1), a deleteriousness threshold suggested by Kircher et al. [32].

Lastly, we assessed the effects of all identified genetic variants on gene regulatory networks in order to prioritize key genes and highlight pathway, tissue, and rare disease phenotype enrichment with Downstreamer [33]. In total, the analysis identified 51 significant key genes (Supplementary Data). For basal cell carcinoma, the key genes showed enrichment in UV-damage excision repair (GO:0070914), protein binding molecular function (GO:0005515), and abnormalities of lymphocyte physiology (HP:0031409), cellular physiology (HP:0011017), cellular phenotype (HP:0025354), of the immune system (HP:0002715, HP:0010978), carious teeth (HP:0000670), and unusual infection (HP:0032101). For breast cancer, the key genes were associated with negative regulation of transcription by RNA polymerase II (GO:0000122) and with abnormalities of the menstrual cycle (HP:0000140, HP:0000858), the digestive system (HP:0025031), oral morphology (HP:0031816, HP:0000163, HP:0006483, HP:0009804, HP:0000164), and of limbs (HP:0040064, HP:0004097, HP:0009484, HP:0001155). For non-Hodgkin’s lymphoma, we found enrichment in telomeric DNA binding function (GO:0042162) and in abnormality of hair pigmentation (HP:0009887). For melanoma, the analysis identified an association with 3 KEGG pathways (allograft rejection, intestinal immune network for IgA production, and autoimmune thyroid disease). For colon cancer, the key genes were enriched also in endometrial cancer and renal cell carcinoma KEGG pathways. To determine how similar the key gene predictions are, we correlated the key gene scores of the 11 cancers to each other. We observed that some gene prioritization scores often have positive correlations, especially between basal cell carcinoma, prostate and breast cancers, highlighting shared genetic signature of the disorders (Figure 3a). Finally, the Downstreamer analysis showed significant enrichment for potentially tumour-associated gene expression in related tissues across 54 GTEx v8 tissues for basal cell carcinoma, breast and prostate cancer S7.

In summary, our novel findings confirm shared genetic architecture between cancers, highlight tumour-associated expression patterns that likely stem from germline variation, and provide additional potential regulatory mechanisms through which germline variation can affect cancer risk.

## Discussion

Our results demonstrate the advantages of using joint Bayesian modelling and age-at-onset phenotypes for genomic prediction and GWAS discovery, highlighting how these approaches can be used to improve the utilisation of existing data. Biobanks of large sample sizes are becoming increasingly common worldwide and improved links to electronic health record data enable information to be obtained regarding age-at-diagnosis. Thus, we expect that our approach of incorporating age-at-onset data in the analysis will become commonplace, improving case-control studies using richer information about the disease process.

One of the fundamental problems of analysing cancer phenotypes in a case-control fashion is the uncertainty that the control group subjects might later be diagnosed with cancer. Many cancers often become more prevalent only at later ages (Figure 2c), and as biobanks primarily consist of young, healthy individuals, it could distort the inference. For example, that issue can be mitigated by introducing age thresholds to eliminate younger individuals who have been at risk only for a limited amount of time or by age-matching individuals. However, this will always be somewhat arbitrary, reducing the sample size, and there is no guarantee that older individuals will not develop cancer later in life. In contrast, time-to-event analysis treats these individuals as right censored, making no additional assumptions about the cancer occurrence in future, which suggests an alternative with a more sound conceptual background to yield more accurate inferences. Interestingly, time-to-event adjustment tends to yield higher power than the case-control adjustment once the case count is sufficiently high. Hence, time-to-event analyses could become more statistically powerful than their case-control counterparts as cases accrue.

There are important caveats to this study. First, our PRS results remain limited as our work represents a re-analysis of a single biobank study to demonstrate the methodological improvements that can be obtained. However, nothing prevents GMRM-BayesW from being run across different studies and posterior mean SNP effects being combined to improve the effectiveness of the PRS, providing predictors with the potential to stratify individuals for screening programs. For example, prostate cancer screening has been found to be only moderately useful for the general population with 17-40% [34, 35] reduction in cancer-specific deaths, but as the mortality rate is low (in USA stage I-III 5-year survival rate >95%, stage IV 5-year survival rate 30% [36]) and as there are potential complications following the treatment, a general screening program has not been commonly implemented. Nevertheless, there are recommendations for stratified risk communication. For example, the American Cancer Society suggests that men with a first-degree relative with prostate cancer before age 65 should be informed about screening and its risks already at age 45, and men with multiple relatives with prostate cancer before 65 should be informed about screening even at age 40 [37]. Moreover, it has been found that even if screening is not cost-effective for men at average risk of prostate cancer, it is still cost-effective for men at very high risk (five times higher risk than the average) [35]. Our results suggest that the top 5% highest Bayesian polygenic risk scores could serve as a rule to detect those who should be screened and whose risk should be communicated.

A second limitation is our study’s restriction to a discovery set of UK Biobank individuals with European ancestry, whereas many other recent studies have instead focused on merging and meta-analysing multiple data sets from various backgrounds. However, our goal was simply to show that discoveries are yet to be made on the existing data set simply by using an enhanced methodology for timing-related traits rather than occurrence-related traits, and these improvements should also transfer when meta-analysing multiple data sets. Third, the low number of cancer cases, such as testicular, ovarian or endometrial cancer, lead to sub-optimal prediction accuracy, while cancers with higher case counts (prostate, breast) yielded larger-scale improvements in prediction accuracy. Again, as stated above, these analyses should be repeated in data with greater case counts for the cancers with smaller case counts. Fourth, our current analysis combines prevalent and incident cases to maximise the statistical power. However, it has been shown [7] that effect sizes are generally very similar even if we restrict the analysis only to incident cases. Future time-to-event analyses could also benefit from using information about left truncation by including the entry date in the analysis, but the gain is likely marginal if the phenotypic information is derived from the medical records and the onset happens later in life.

In summary, we have shown random effect models which utilise time-to-event data, maximise the use of existing data for 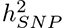 estimation, genomic prediction and GWAS discovery of 11 common cancers.

## Methods

### UK Biobank Data

We restricted our discovery analysis in the UK Biobank to a sample of European-ancestry individuals. To infer ancestry, we used both self-reported ethnic background (UK Biobank field 21000-0), selecting coding 1, and genetic ethnicity (UK Biobank field 22006-0), selecting coding 1. We also took the 488,377 genotyped participants and projected them onto the first two genotypic principal components (PC) calculated from 2,504 individuals of the 1,000 Genomes project with known ancestries. Using the obtained PC loadings, we then assigned each participant to the closest population in the 1000 Genomes data: European, African, East-Asian, South-Asian or Admixed, selecting individuals with PC1 projection < absolute value 4 and PC 2 projection < absolute value 3. Samples were also excluded if in the UK Biobank quality control procedures they (i) were identified as extreme heterozygosity or missing genotype outliers; (ii) had a genetically inferred gender that did not match the self-reported gender; (iii) were identified to have putative sex chromosome aneuploidy; (iv) were excluded from kinship inference; (v) had withdrawn their consent for their data to be used. We used genotype probabilities from version 3 of the imputed autosomal genotype data provided by the UK Biobank to hard-call the genotypes for variants with an imputation quality score above 0.3. The hard-call-threshold was 0.1, setting the genotypes with probability ≤ 0.9 as missing. From the good quality markers (with missingness less than 5% and p-value for Hardy-Weinberg test larger than 10^−6^, as determined in the set of unrelated Europeans) we selected those with minor allele frequency (MAF) > 0.0002 and rs identifier, in the set of European-ancestry participants, providing a data set 9,144,511 SNPs. From this, we took the overlap with the Estonian Biobank data described below to give a final set of 8,430,446 markers. This provides a set of high quality SNP markers present across both discovery and prediction data sets. For computational convenience when conducting the joint Bayesian analysis we created an additional subset of markers by removing markers in very high LD, through the selection of the highest MAF marker from any set of markers with LD *R*^2^ ≥ 0.8 within a 1Mb window. These filters resulted in a data set with 458,747 individuals and 2,174,071 markers.

We used the recorded measures of individuals to generate the phenotypic data sets for 11 most common types of cancer: bladder, breast, cervix, colon, endometrium, ovary, prostate, testis, basal cell carcinoma, melanoma, and non-Hodgkin’s lymphoma. Then, we created time-to-event phenotypes using either self-reported data or the linked electronic medical records data. For the medical record data, we used UK Biobank field 40008 to get the earliest age at each cancer diagnosis together with fields 40006 and 40013 to indicate the ICD10 or ICD9 cancer type (Table S2). Individuals without an entry on those fields were considered censored and their time was set to their last known age alive (exact birth date imputed to day 15 of a month as only month and year are known) without a cancer diagnosis. Each individual *i* was therefore assigned a censoring indicator *C_i_* that was defined *C_i_* = 1 if the person had the event before the end of the follow-up period and *C_i_* = 0 otherwise. For self-reported time-to-event phenotypes, we created a pair of last known time (averaged between assessments) without an event and censoring indicator *C_i_*. Similarly to the medical record phenotypes, if the event had not happened, then the last time without having the event was defined as the last date of assessment centre or date of death visit minus date of birth. For creating the self-reported phenotypes, we used UK Biobank field 20001 for the presence or absence of certain cancer type and UK Biobank field 20007 for interpolated ages of individuals when the disease was first diagnosed; we excluded from the self-reported phenotype analysis individuals who said that they had cancer, but there was no record of the diagnosis age. In an attempt to further increase power and to account for potential missingness, for each individual who had self-reported data about cancer timing but no medical record data, we used used the self-reported age-at-diagnosis instead of treating the individual as censored. However, this approach only yielded marginal improvements compared to using purely medical record information (Figure S4). Finally, the case-control-phenotypes corresponding to the time-to-event phenotypes were defined as the censoring indicators *C_i_*.

The analyses were adjusted for the following covariates: sex for sex-unspecific cancers, age in case-control analyses, UK Biobank recruitment center, home location, genotype batch and 20 first principal components (UK Biobank field 22009) to account for the population stratification in a standard way. For the analyses that used age-at-diagnosis as phenotypes, we did not include any covariates of age or year of birth because these are directly associated to our phenotypes.

### Estonian Biobank Data

For the Estonian Biobank data, 195,432 individuals were genotyped on Illumina Global Screening (GSA) arrays and we imputed the data set to an Estonian reference, created from the whole genome sequence data of 2,244 participants [38]. We kept only the European ancestry individuals with available information on sex, age-at-recruitment and date of recruitment. From 11,130,313 markers with imputation quality score > 0.3, we selected SNPs that overlapped with those selected in the UK Biobank, resulting in a set of 8,430,446 variants out of which 2,174,071 variants were used in the prediction analysis. The 60 previously unreported variants that were found significant in the marginal association analysis of UK Biobank (Table S5) were used in a replication analysis using the same Estonian Biobank individuals.

We created the phenotypes for the Estonian Biobank individuals using the respective medical record information. The occurrence of each of the cancers was defined by using the respective ICD10 codes exactly as it was defined for the UK Biobank medical record phenotypes (Supplementary table S2) by first defining the last known time person did not have a respective diagnosis. Individuals with a respective cancer diagnosis received a censoring indicator *C_i_* = 1 and 0 otherwise that then defined the case-control phenotypes adjusted for covariates such as sex for sex-unspecific cancers and age.

### Analysis with joint Bayesian models

Both GMRM-BayesRR-RC and GMRM-BayesW models are based on grouped effects with spike-and-slab mixture priors and provide joint effect estimates [20, 39]. Briefly, genetic markers are grouped into annotation-specific sets, e.g. based on MAF or LD, with independent hyperparameters for the phenotypic variance attributable to each group. This allows estimation of the phenotypic variance attributable to the group-specific effects.

Assuming *N* individuals, *M* genetic markers, and Φ groups, the GMRM-BayesRR-RC model of an observed phenotype vector **y** is:

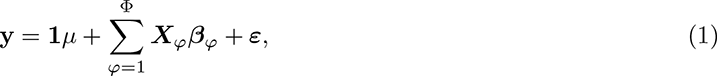

where ***X****_ϕ_*is a standardised genotype matrix containing SNPs allocated to group *ϕ*, *µ* is an intercept, ***β****_ϕ_*is the vector of SNP effects in group *ϕ* and ***ε*** is a vector of Gaussian residuals such that every element in the vector would be distributed as 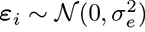. For each group, we assume that ***β****_ϕ_* are distributed according to a mixture of Gaussian components with mixture specific proportions ***π****_ϕ_* and mixture variances 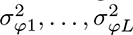 and a Dirac delta at zero which induces sparsity:

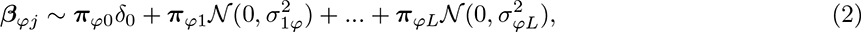

where *L* is the number of mixture components.

The GMRM-BayesW follows similar prior and grouping formulation. However, here we assume for an individual *i* that the age-at-onset of a disease *y_i_* has Weibull distribution, with a reparametrisation of the model to represent the mean and the variance of the logarithm of the phenotype as

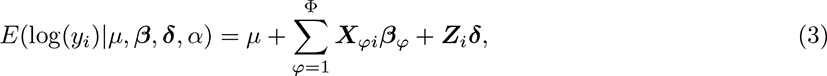

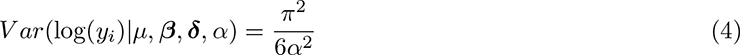

where ***Z****_i_* are additional covariates (such as sex or genetic principal components), ***δ*** are the additional covariate effect estimates and *α* is the Weibull shape parameter.

We estimated the hyperparameters such as genetic variance and prior inclusion probability by grouping markers into MAF-LD bins as recent theory suggests this yields improved estimation [39–41]. We ran the GMRM-BayesW model on the UK Biobank data with 8 MAF-LD groups that were defined as first splitting markers by MAF quartiles and then splitting each of those MAF quartiles into two LD score blocks (MAF quartiles are 0.007, 0.020, 0.102; median LD score in each quartile are 2.32, 3.33, 5.69, 9.25, from the lowest MAF quartile respectively). We decided not to split these groups further as the potentially low statistical power of cancer-related phenotypes could lead to many groups with zero genetic variance. Both GMRM-BayesW and GMRM-BayesRR-RC models were executed with mixture components (0.0001, 0.001, 0.01, 0.1) for each of the groups, reflecting that the markers allocated into those mixtures explain the magnitude of 0.01%, 0.1%, 1% or 10% of the group-specific genetic variance. We ran the GMRM-BayesW model using the timing of cancers as the phenotype while treating individuals without cancer as censored, and we ran the GMRM-BayesRR-RC type model using the occurrence of cancer as the phenotype. In the GMRM-BayesW analyses we took the covariates into account by estimating the effects of the covariates within the GMRM-BayesW model while in the GMRM-BayesRR-RC we regressed out the covariates from the phenotype prior to the analysis.

We specified the hyperparameters for the models such that they would be weakly informative. For GMRM-BayesW model, the choice of hyperparameters and quadrature points was exactly the same as in [20]; for GMRM-BayesRR-RC model the choice was exactly the same in [39]. We ran the chains for each of the cancer types twice for 6000 iterations, discarding the first 2000 iterations as a burn-in and using a thinning step of 5, leaving us with a final of 1600 samples of the posterior distribution. As estimation is done in parallel, the run time will depend on the degree of parallelisation. For example, for basal cell carcinoma we used 11 nodes and 12 tasks per node (total 132 tasks) for GMRM-BayesW and 7 nodes and 12 tasks per node (total 84 tasks) for GMRM-BayesRR-RC. This resulted in a total run time of 67.5 hours (40.5s per iteration) for GMRM-BayesW and 79.7 hours (47.8s per iteration) for GMRM-BayesRR-RC. Although GMRM-BayesW was faster in the absolute time, adjusted for the number of tasks in the example, GMRM-BayesW requires 33% more time per iteration than GMRM-BayesRR-RC. Other choices for parallelisation (for example synchronisation rate) were set the same as described in [20] and note that these timings were recorded prior to the recent speed-ups in our GMRM-BayesRR-RC model [15].

### Association testing with adjusted marginal models

Since many of the assessed cancer traits had imbalanced case-control ratios, we adapted single-variant association testing of REGENIE [17], a two-step method designed to control Type I error in GWAS by using Firth logistic regression or Saddle Point Approximation (SPA). However, instead of running the first step of REGENIE, where 23 leave one chromosome out (LOCO) genetic predictors are generated based on stacked block Ridge regression, we provided the LOCO predictors from the either GMRM-BayesRR-RC or GMRM-BayesW joint analysis:

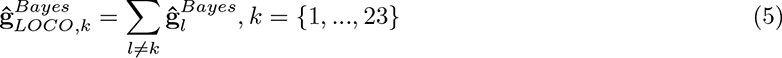

Here, 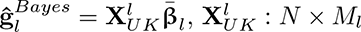 matrix of SNPs in the *l*th chromosome, 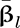 is the vector of average effect sizes from joint Bayesian analysis in chromosome *l*. These Bayesian LOCO predictors were used in the second step of REGENIE that tests genetic markers for association with the phenotype conditional upon the random polygenic effects from other chromosomes 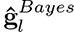:

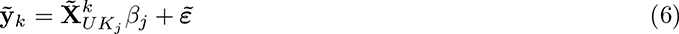

where 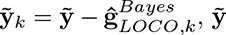 is the binary phenotype, 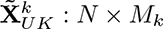 matrix of SNPs in the *k*th chromosome, 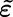 is the error term, *β_j_* is the *j*th SNP effect that we are estimating. All tilde symbols represent adjustment for covariates. We refer to this modified Bayesian version of REGENIE analysis as "GMRM-BayesRR-RC-adjusted" or "GMRM-BayesW-adjusted".

For comparison, we ran the standard REGENIE pipeline (Equation 6) with LOCO predictors from step 1 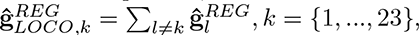 to which we refer as "unadjusted" analysis.

We used the full overlap of UK Biobank and Estonian Biobank markers giving us a total of 8,433,421 markers to be analysed. We provided sex, (if applicable for the cancer), age, UK Biobank assessment centre, coordinates of birthplace, genotype chip, and the leading 20 PCs of the SNP data as covariates. In the first step of REGENIE, we followed recommendations in the [17] and excluded SNPs in inter-chromosomal LD with *R*^2^ *>* 0.1147 from the unadjusted analysis. In the second step of REGENIE, we ran association testing with approximate Firth correction. However, we encountered convergence issues for all 4 female cancers and Testicular cancer potentially due to a very low case count and thus used SPA instead of the Firth correction for these 5 traits in both adjusted and unadjusted analyses. We used the p-value threshold of 5 · 10^−8^ to determine the significance of each marker.

We applied the following steps to the association results to filter out independent and potentially previously undiscovered markers. Firstly, we LD clumped the results such that the index SNPs would have a p-value below 5 · 10^−8^ and SNPs could be added to a clump if they were 1Mb from the index SNP, they were correlated with *r*^2^ *>* 0.05 and they were nominally significant (*P <* 0.05). Next, we used COJO method [42] from GCTA software [43] to find clumps with independent signals by conducting a stepwise selection of index SNPs in a 1Mb window and we considered SNPs independent if they had a p-value below 5 · 10^−8^ in the joint model. To determine novelty, we first removed all markers that were significantly associated in the unadjusted model. We then removed all the markers that had a correlation of *r*^2^ *>* 0.1 with a marker that had been previously found associated with a cancer of interest using GWAS Catalog (published until April 2022) and the LDtrait tool with the British in England and Scotland population. We then again used COJO to condition the remaining markers on the previously identified associations for each cancer of interest and SNPs that did not fall below 5 · 10^−8^ in the joint model were eliminated. For the remaining SNPs we conducted an additional literature review using the Phenoscanner database [44, 45] to find any previous associations with variants of interest or variants in LD. The remaining candidates of novel associations were concatenated across GMRM-BayesW or GMRM-BayesRR-RC adjusted analyses and then included in the replication analysis using the largest available studies conducted for each specific cancer type. We checked our findings for replication in the FinnGen summary statistics version R8 [46]. Replication was defined as Bonferroni corrected p-value being lower than 0.05 and the direction of the effect size same in both the original analysis and the replication analysis.

### Liability scale heritability and genetic correlation

We used the summary statistics from the marginal association analysis in LD score regression [23] to calculate the observed scale heritability. We used the LD scores from the 1000 Genomes European data https://alkesgroup.broadinstitute.org/LDSCORE/ and the summary statistics were taken from either GMRM-BayesW-adjusted or GMRM-BayesRR-RC-adjusted association analysis. The conversion of the heritability to the liability scale was done using the formula by Lee et al. [47] (Table 2) and using the risks from SEER 2016-2018 (Table S6) [48] using the risks of having cancer diagnoses between ages 0 to 85 for non-Hispanic white people providing similarity with the study population (European ancestry, UK Biobank, oldest person age 86). We further provide an alternative liability scale transformation [24] designed for rare traits. Using the alternative rare trait liability scale transformation we also present the heritability estimates from the joint Bayesian case-control model (Table S3). In addition, we used cross-trait LD score regression [49] to calculate the genetic correlations using results from REGENIE’s step 2 analyses with GMRM-BayesW or GMRM-BayesRR-RC adjustments and GWAS results for multiple phenotypes released by Neale group [50] and Global Biobank Meta-analysis Initiative consortium [51]. The significance threshold of 0.05 was corrected by the total number of tests (2200).

**Table 2.** SNP-heritability estimates. Estimates with 95% CI from LD Score regression, using mixed linear association model estimates from REGENIE’s step 2 adjusted with age-at-onset LOCO predictor (GMRM-BayesW) or with case-control LOCO predictor (GMRM-BayesRR-RC), as compared with previous array or family based estimates. a - estimate from Rashkin et al. [7]; b - estimate from Mucci et al. [59]; c - estimate from Kilgour et al. [60]; d - estimates from Czene et al. [61].

|  | BayesW | BayesRR-RC | Array-based estimate<sup>a</sup> | Family-based estimate<sup>b</sup> |
| --- | --- | --- | --- | --- |
| Basal cell carcinoma | 0.56 (0.36-0.76) | 0.56 (0.36-0.76) | 0.17 (0.07-0.27)<sup>c</sup> | 0.43 (0.26-0.59) |
| Bladder cancer | 0.06 (0-0.13) | 0.06 (0-0.12) | 0.08 (0.04-0.12) | 0.07 (0.02-0.11) |
| Breast cancer | 0.20 (0.13-0.27) | 0.20 (0.13-0.27) | 0.10 (0.08-0.13) | 0.31 (0.11-0.51) |
| Cervical cancer | 0.05 (0.03-0.06) | 0.05 (0.03-0.06) | 0.07 (0.02-0.12) | 0.13 (0.06-0.15)<sup>d</sup> |
| Colon cancer | 0.08 (0.04-0.13) | 0.08 (0.04-0.13) | 0.07 (0.04-0.10) | 0.15 (0.00-0.45) |
| Endometrial cancer | 0.07 (0-0.16) | 0.07 (0-0.15) | 0.13 (0.07-0.18) | 0.27 (0.11-0.43) |
| Melanoma | 0.15 (0.03-0.27) | 0.15 (0.03-0.27) | 0.08 (0.04-0.11) | 0.58 (0.43-0.73) |
| Non-Hodgkin's lymphoma | 0.04 (0-0.10) | 0.04 (0-0.10) | 0.13 (0.03-0.23) | 0.10 (0.08-0.10)<sup>d</sup> |
| Ovarian cancer | 0.01 (0-0.10) | 0.01 (0-0.10) | 0.07 (0.01-0.13) | 0.39 (0.23-0.55) |
| Prostate cancer | 0.48 (0.21-0.76) | 0.48 (0.2-0.76) | 0.16 (0.13-0.20) | 0.57 (0.51-0.63) |
| Testicular cancer | 0.56 (0.03-1.08) | 0.55 (0.02-1.08) | 0.26 (0.15-0.38) | 0.37 (0.00-0.93) |

### Discovery follow-up analyses

We conducted a number of follow-up analyses using the marginal summary statistics obtained from the mixed-linear association model REGENIE that was adjusted with GMRM-BayesW and GMRM-BayesRR-RC LOCO predictors instead of built-in ridge LOCO predictors.

We used FUMA (Functional Mapping and Annotation) [52] platform to functionally characterise novel replicated variants and prioritise genes. We defined a threshold for independent significant novel SNPs and corresponding novel genetic regions as LD *r*^2^ = 0.6 on the reference panel UKB/release2b. When performing gene mapping, we used 10kb maximum distance for positional mapping, all available tissue types and maximum p-value threshold of 5 · 10^−8^ for eQTL mapping, and builtin chromatin interaction data with 1 · 10^−6^ FDR threshold for chromatin interaction mapping. We obtained ANNOVAR’s functional annotation and nearest genes [52, 53], RegulomeDB categorical score [26], eQTL information, 15-core chromatin state [27], and CADD deleteriousness score [32] from SNP2GENE analysis of FUMA [52]. We considered markers as deleterious if their CADD score exceeded the 12.37 threshold suggested by Kircher et al. (2014) [32] and chromatin state as open/active for SNPs with 15-core chromatin score ≤7 predicted by ChromHMM [27] based on 5 chromatin marks for 127 epigenomes.

We annotated the SNPs with DeepSEA Beluga and DeepSEA Sei, deep-learning-based frameworks for systematical prediction of the chromatin effects of sequence alterations and sequence regulatory activities, respectively [28, 29]. For each novel genomic region, we report here the DeepSEA Beluga’s maximum mean log e-value (MLE) and disease impact score (DIS) and Sei’s sequence class with the maximum absolute score. We calculated all of the mentioned parameters for each significant independent novel SNP as well as minimum/maximum/common values within the novel genetic regions.

We investigated the effects of identified genetic variants on gene regulatory networks by using Downstreamer [33] with the GMRM-BayesW and GMRM-BayesRR-RC marginal summary statistics. This method helped to prioritize key genes and highlight pathway, tissue, and rare disease phenotype enrichment. For key gene enrichment score correlation, we used both significant and insignificant enrichment Z-scores. For the rest of the analysis, we used those genes/samples/pathways/GO terms/tissues which passed both Bonferroni and 5% FDR significance thresholds.

### Genomic prediction in the Estonian Biobank

The predictors based on GMRM-BayesW or GMRM-BayesRR-RC models into Estonian Biobank 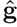 were obtained by multiplying the standardised genotype matrix with the average SNP effect across iterations

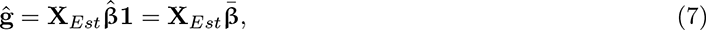

where **X***_Est_* is *N_Est_ × M* matrix of standardised Estonian genotypes (each column is standardised using the mean and the standard deviation of the Estonian data), 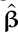 is the *M × I* matrix containing the posterior distributions for *M* marker effect sizes across *I* iterations, 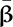 is the average SNP effect. We calculated the average predictor from GMRM-BayesW and GMRM-BayesRR-RC models for each cancer using 1600 iterations (see Analysis with joint Bayesian models). To facilitate comparison between marginal and joint estimates, we used the same training data set (same 2,174,071 variants and 458,747 individuals) from UK Biobank to estimate the marginal summary statistics for the 11 cancers. The summary statistics were estimated using fastGWA [14], taking into account the relatedness in the data. Then, we used these summary statistics in LDpred-funct method [25]. As annotations for LDpred-funct, we used 8 MAF-LD bins similarly to the Bayesian analyses; we used an LD radius of 1Mb; heritability estimates were taken from LDscore regression, using the fastGWA summary statistics. The weights from LDpred-funct were then applied to the Estonian Biobank individuals to create the respective genomic predictors that were compared with the GMRM-BayesW and GMRM-BayesRR-RC results.

We evaluated the performance of the three genetic predictors for each cancer phenotype by comparing them to the actual phenotype case-control status using logistic regression and true phenotype timing using Cox proportional hazards (PH) model. The three predictors were compared using the top 5% PRS with the rest, the top 10% PRS with the rest, and the effect of one standard deviation increase in PRS. From the logistic regression, we calculated odds ratios for the top 5%, top 10% and scaled change effect. From the Cox PH model, we calculated hazard ratios and Harrell’s C-statistics [54] for the top 5%, top 10% and scaled change effect. In addition to the predictor, gender (if applicable) and age-at-entry were included in the logistic regression and Cox PH model that calculated the hazard ratio. The Cox PH model calculated Harrell’s C-statistic, where the true phenotype was regressed only on the predictor. The results of odds ratios, hazards ratios and Harrell’s C-statistics are shown in Figure 2a and Supplementary figure S3. We further used the top 5% and top 10% PRS individuals to see what percentage of them develop cancer (Figure 2b).

Across all the cancers and 4 predictive scores we calculated the respective cumulative incidence curves for the top 5% highest PRS individuals (Figure 2c) adjusting the analysis for the competing risks. The calculation was done using R package cmprsk [55, 56].

## Data Availability

All data produced in the present study are available upon reasonable request to the authors.

## Author contributions

SEO and MRR conceived and designed the study. MRR, SEO, ESM designed the study with contributions from KL and MCS. SEO, ESM, KL, MCS and MRR contributed to the analysis. SEO, ESM, MCS and MRR wrote the paper. KL, ZK, RM provided study oversight, contributed data and ran computer code for the analysis. All authors approved the final manuscript prior to submission.

## Data availability

This project uses UK Biobank data under project 35520. UK Biobank genotypic and phenotypic data is available through a formal request at (http://www.ukbiobank.ac.uk). The UK Biobank has ethics approval from the North West Multi-centre Research Ethics Committee (MREC). For access to be granted to the Estonian Biobank genotypic and corresponding phenotypic data, a preliminary application must be presented to the oversight committee, who must first approve the project, ethics permission must then be obtained from the Estonian Committee on Bioethics and Human Research, and finally a full project must be submitted and approved by the Estonian Biobank. This project was granted ethics approval by the Estonian Committee on Bioethics and Human Research (https://genomics.ut.ee/en/biobank.ee/data-access). The activities of the EstBB are regulated by the Human Genes Research Act, which was adopted in 2000 specifically for the operations of the EstBB. Individual level data analysis in the EstBB was carried out under ethical approval 1.1-12/624 from the Estonian Committee on Bioethics and Human Research (Estonian Ministry of Social Affairs), using data according to release application S16 from the Estonian Biobank.

## Code availability

The BayesW model was executed with the software Hydra, with full open source code available at https://github.com/medical-genomics-group/hydra [57]. Summary MR-HEIDI tests were conducted using the SMR software (version 1.03) [58]. The multivariable MR analyses were carried out with SMR-IVW extension software https://github.com/masadler/smrivw. plink version 1.9 is available at https://www.cog-genomics.org/plink/. R version 4.0.3 is available at https://www.r-project.org/.

## Acknowledgements

This project was funded by an SNSF Eccellenza Grant to MRR (PCEGP3-181181), and by core funding from the Institute of Science and Technology Austria and the University of Lausanne. K.L. and R.M. were supported by the Estonian Research Council grant PRG687. Estonian Biobank computations were performed in the High Performance Computing Centre, University of Tartu. We would like to acknowledge the participants and investigators of UK Biobank, FinnGen and Estonian Biobank studies. This project uses UK Biobank data under project number 35520.

## Supplementary material

### Supplementary tables

**Table S1.** UK Biobank data composition for the cancer cases and their timings used within the study.

|  | All |  |  | Females |  |  | Males |  |  |
| --- | --- | --- | --- | --- | --- | --- | --- | --- | --- |
|  | Cases (%) | Mean (sd) | Median (Range) | Cases (%) | Mean (sd) | Median (Range) | Cases (%) | Mean (sd) | Median (Range) |
| Basal cell carcinoma | 26758 (5.8%) | 60.6 (9.42) | 62.1 (7.5-80.6) | 13277 (5.3%) | 59.5 (9.73) | 61 (7.5-80.6) | 13481 (6.4%) | 61.8 (8.96) | 63.3 (22.2-79.7) |
| Bladder cancer | 2470 (0.5%) | 61.3 (9.38) | 62.9 (19.1-77.8) | 608 (0.2%) | 60.8 (9.50) | 62.1 (24.7-76.9) | 1862 (0.9%) | 61.5 (9.34) | 63.1 (19.1-77.8) |
| Breast cancer | 16972 (6.8%) | 55.7 (9.20) | 55.6 (18.8-80.9) | 16972 (6.8%) | 55.7 (9.20) | 55.6 (18.8-80.9) |  |  |  |
| Cervical cancer | 8680 (3.5%) | 38.0 (9.27) | 36.8 (13.5-76.6) | 8680 (3.5%) | 38.0 (9.27) | 36.8 (13.5-76.6) |  |  |  |
| Colon cancer | 4463 (1.0%) | 60.9 (9.13) | 62.3 (11.2-78.8) | 2009 (0.8%) | 60.2 (9.53) | 61.4 (11.2-78.0) | 2454 (1.2%) | 61.5 (8.76) | 62.7 (14.0-78.8) |
| Endometrial cancer | 2227 (0.9%) | 56.5 (11.22) | 58.3 (13.0-76.9) | 2227 (0.9%) | 56.5 (11.22) | 58.3 (13.0-76.9) |  |  |  |
| Melanoma | 5778 (1.3%) | 54.1 (12.16) | 55.8 (0.5-77.5) | 3243 (1.3%) | 52.1 (12.36) | 53.3 (1.5-77.5) | 2535 (1.2%) | 56.6 (11.41) | 58.5 (0.5-77.5) |
| Non-Hodgkin's lymphoma | 2298 (0.5%) | 58.7 (11.36) | 61.0 (3.5-79.1) | 1026 (0.4%) | 58.8 (10.97) | 60.9 (5.3-78.1) | 1272 (0.6%) | 58.7 (11.67) | 61.0 (3.5-79.1) |
| Ovarian cancer | 1573 (0.6%) | 54.6 (12.38) | 56.0 (10.1-79.6) | 1573 (0.6%) | 54.6 (12.38) | 56.0 (10.1-79.6) |  |  |  |
| Prostate cancer | 9824 (4.7%) | 64.6 (5.87) | 65.2 (22.9-80.2) |  |  |  | 9824 (4.7%) | 64.6 (5.87) | 65.2 (22.9-80.2) |
| Testicular cancer | 886 (0.4%) | 40.5 (11.08) | 40.0 (0.5-76.0) |  |  |  | 886 (0.4%) | 40.5 (11.08) | 40.0 (0.5-76.0) |

**Table S2.** Cancer-specific ICD10 and ICD9 codes used to select cases from the UK and Estonian biobank studies. For each of the tumour types, the corresponding ICD10 and ICD9 codes are presented that were used to define cancer occurrence.

|  | ICD10 code | ICD9 code |
| --- | --- | --- |
| Basal cell carcinoma | C44.0, C44.1, C44.2, C44.3, C44.4, C44.5, C44.6, C44.7, C44.8, C44.9, D04.0, D04.1, D04.2, D04.3, D04.4, D04.5, D04.6, D04.7, D04.8, D04.9 | 173.0, 173.1, 173.2, 173.3, 173.4, 173.5, 173.6, 173.7, 173.8, 173.9, 232.1, 232.2, 232.3, 232.4, 232.5, 232.6, 232.7, 232.8, 232.9 |
| Bladder cancer | C67.0, C67.1, C67.2, C67.3, C67.4, C67.5, C67.6, C67.6, C67.7, C67.8, C67.9, D09.0 | 188.0, 188.2, 188.4, 188.6, 188.8, 188.9, 233.7 |
| Breast cancer | C50.0, C50.1, C50.2, C50.3, C50.4, C50.5, C50.6, C50.7, C50.8, C50.9, D05.0, D05.1, D05.7, D05.9 | 174.0, 174.1, 174.2, 174.3, 174.4, 174.5, 174.6, 174.7, 174.8, 174.9, 233.0 |
| Cervical cancer | C53.0, C53.1, C53.8, C53.9, D06.0, D06.1, D06.7, D06.9 | 180.0, 180.1, 180.8, 180.9, 233.1 |
| Colon cancer | C18.0, C18.1, C18.2, C18.3, C18.4, C18.5, C18.6, C18.7, C18.8, C18.9, D01.0 | 153.0, 153.1, 153.2, 153.3, 153.4, 153.5, 153.6, 153.7, 153.8, 153.9, 230.3 |
| Endometrial cancer | C54.1, D07.0 | 182.0 |
| Melanoma | C43.0, C43.1, C43.2, C43.3, C43.4, C43.5, C43.6, C43.7, C43.8, C43.9 | 172.0, 172.1, 172.2, 172.3, 172.4, 172.5, 172.6, 172.7, 172.8, 172.9 |
| Non-Hodgkin's lymphoma | C82.0, C82.1, C82.2, C82.7, C82.9, C83.0, C83.1, C83.2, C83.3, C83.4, C83.5, C83.6, C83.7, C83.8, C83.9, C85.0, C85.1, C85.7, C85.9 | 202.8 |
| Ovarian cancer | C56 | 183.0 |
| Prostate cancer | C61, D07.5 | 185 |
| Testicular cancer | C62.0, C62.1, C62.9 | 186.9 |

**Table S3.** Alternative liability scale heritability estimates with 95% CI. We use the observed scale from LDSC estimates (REGENIE’s summary statistics from GMRM-BayesW-adjusted analysis) and the heritability estimates from the full Bayesian model (GMRM-BayesRR-RC). Transformation of the observed scale heritabilities is done with a more conservative approach (Ojavee et al. [24]) better suited for rare diseases.

|  | Alternative LDSC<br>GMRM-BayesW-adjusted | Full Bayesian<br>GMRM-BayesR |
| --- | --- | --- |
| Basal cell carcinoma | 0.33 (0.18-0.48) | 0.22 (0.2-0.24) |
| Bladder cancer | 0.04 (0-0.1) | 0.29 (0.19-0.42) |
| Breast cancer | 0.16 (0.09-0.24) | 0.2 (0.17-0.23) |
| Cervical cancer | 0.08 (0.04-0.11) | 0.13 (0.08-0.2) |
| Colon cancer | 0.07 (0.03-0.11) | 0.19 (0.12-0.28) |
| Endometrial cancer | 0.05 (0-0.12) | 0.34 (0.23-0.47) |
| Melanoma | 0.12 (0.01-0.23) | 0.18 (0.13-0.24) |
| Non-Hodgkin's lymphoma | 0.03 (0-0.08) | 0.32 (0.23-0.43) |
| Ovarian cancer | 0.01 (0-0.09) | 0.49 (0.37-0.61) |
| Prostate cancer | 0.35 (0.1-0.61) | 0.33 (0.28-0.4) |
| Testicular cancer | 0.5 (0-1.05) | 0.81 (0.73-0.87) |

**Table S4.** Statistically significant cross-trait genetic correlations from LD score regression analysis. We calculated the genetic correlations between cancers with cross-trait LD score regression [49] applying it to the results from REGENIE’s GMRM-BayesW or GMRM-BayesRR-RC adjusted analyses and GWAS results for multiple phenotypes released by Neale group [50] and Global Biobank Meta-analysis Initiative consortium [51]. Both GMRM-BayesRR-RC and GMRM-BayesW based significant genetic correlations agree on the magnitude of the estimates.

| Trait 1 | Trait 2 | BayesW (95% CI) | BayesRR-RC (95% CI) |
| --- | --- | --- | --- |
| Basal Cell Carcinoma | BMI | -0.08 (-0.11, -0.05) | -0.08 (-0.11, -0.05) |
| Basal Cell Carcinoma | Hypothyroidism | -0.13 (-0.18, -0.08) | -0.13 (-0.18, -0.08) |
| Cervical Cancer | Age First Live Birth | -0.4 (-0.55, -0.24) | -0.38 (-0.54, -0.23) |
| Cervical Cancer | Age Last Live Birth | -0.46 (-0.64, -0.28) | -0.45 (-0.62, -0.27) |
| Cervical Cancer | Age Started Oral Contraceptive | -0.43 (-0.62, -0.23) | -0.42 (-0.61, -0.22) |
| Cervical Cancer | Educational Attainment | -0.27 (-0.38, -0.17) | -0.26 (-0.36, -0.16) |
| Cervical Cancer | Ever Smoked | 0.29 (0.17, 0.41) | 0.28 (0.16, 0.4) |
| Cervical Cancer | Maternal Smoking Around Birth | 0.31 (0.18, 0.44) | 0.29 (0.17, 0.42) |
| Cervical Cancer | Smoking Status | -0.4 (-0.53, -0.28) | -0.39 (-0.52, -0.27) |
| Melanoma | Basal Cell Carcinoma | 0.44 (0.32, 0.57) | 0.44 (0.32, 0.57) |

**Table S5.**
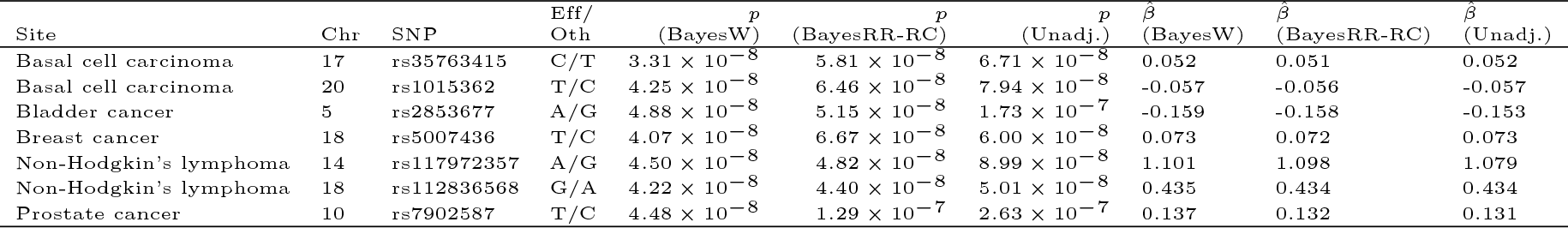
Previously unreported discoveries from GMRM-BayesRR-RC or GMRM-BayesW analyses in comparison with results from an unadjusted marginal association analysis. We observe that for the 7 previously unreported variants, the p-value in the unadjusted association analysis with REGENIE is borderline significant (5 · 10^−8^ *< p <* 10^−6^). However, by using the GMRM-BayesW or GMRM-BayesRR-RC adjustments in the step 1 of REGENIE, we arrive at statistically significant test statistics.

**Table S6.**
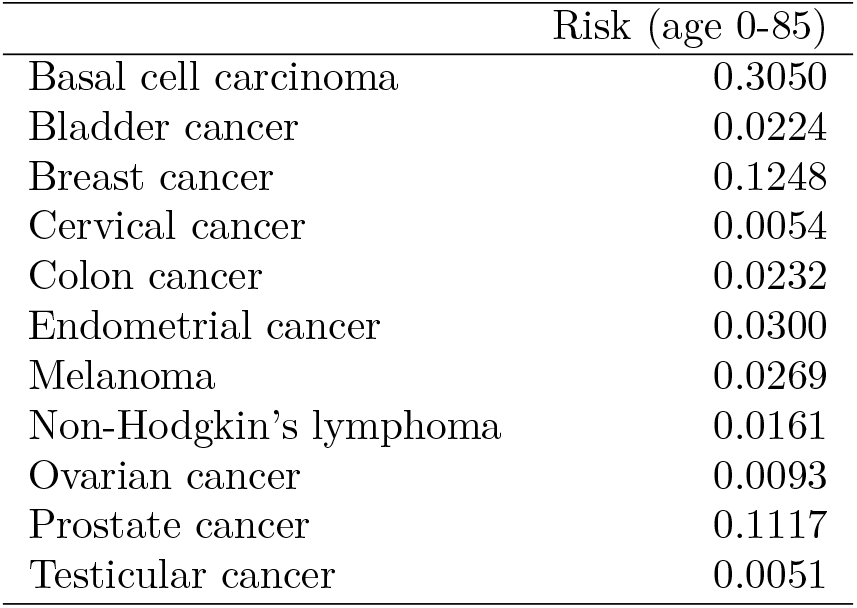
Cancer risk from birth to age 85, SEER estimate 2016-2018. To ensure that the lifetime risk estimates were similar to the study population (European ancestry, UK Biobank, oldest individual age 86) we used the estimates from SEER of non-hispanic white of getting diagnosed between ages (0-85). The explorer is accessible from https://seer.cancer.gov/explorer/. The explorer had a joint estimate for colorectal cancer that we transformed to the risk of colon cancer using the proportion of colon cancer cases among colorectal cases (70.3%, https://www.cancer.org/cancer/colon-rectal-cancer/about/key-statistics.html, accessed 24.01.2022). For basal cell carcinoma, we used a lifetime risk estimate from Miller et al. [62].

|  | Risk (age 0-85) |
| --- | --- |
| Basal cell carcinoma | 0.3050 |
| Bladder cancer | 0.0224 |
| Breast cancer | 0.1248 |
| Cervical cancer | 0.0054 |
| Colon cancer | 0.0232 |
| Endometrial cancer | 0.0300 |
| Melanoma | 0.0269 |
| Non-Hodgkin's lymphoma | 0.0161 |
| Ovarian cancer | 0.0093 |
| Prostate cancer | 0.1117 |
| Testicular cancer | 0.0051 |

### Supplementary figures

**Figure S1.**
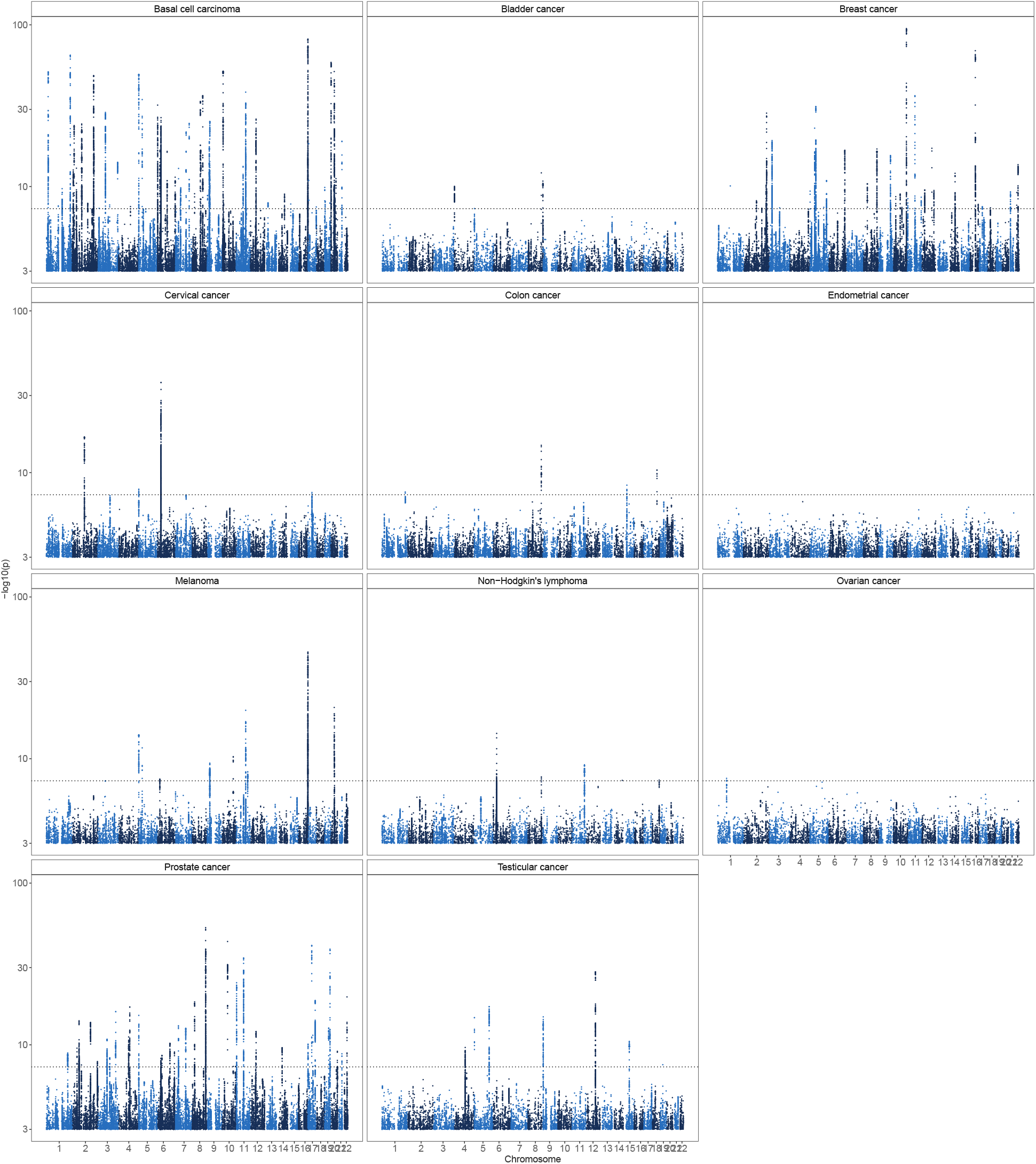
Results from case-control association analysis of 11 tumours, adjusted for BayesW predictors in other chromosomes. The significance of each SNP was obtained using a logistic regression score test from step 2 of REGENIE on binary (case-control) phenotype that was adjusted for covariates and BayesW genetic LOCO predictor. The number of markers analysed was *M* =8,430,446, the number of individuals and cases for each specific cancer are shown in the Supplementary information. We present the − log_10_(p-value), the dotted line indicates a significance threshold of *p* = 5 · 10^−8^.

**Figure S2.**
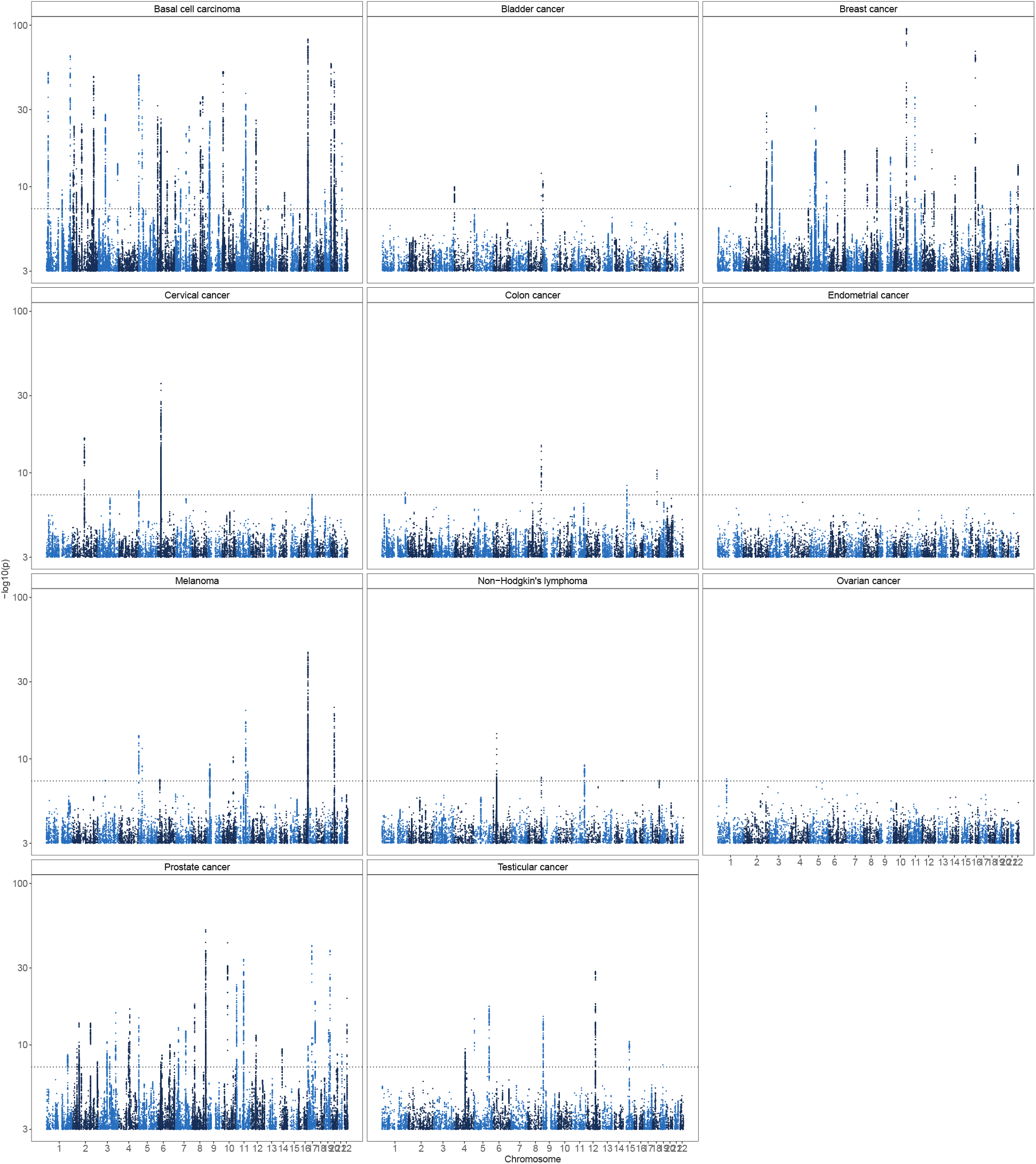
Results from case-control association analysis of 11 tumours, adjusted for BayesRR-RC predictors in other chromosomes. The significance of each SNP was obtained using a logistic regression score test from step 2 of REGENIE on binary (case-control) phenotype that was adjusted for covariates and BayesRR-RC genetic LOCO predictor. The number of markers analysed was *M* =8,430,446, the number of individuals and cases for each specific cancer are shown in the Supplementary information. We present the − log_10_(p-value), the dotted line indicates a significance threshold of *p* = 5 · 10^−8^.

**Figure S3.**
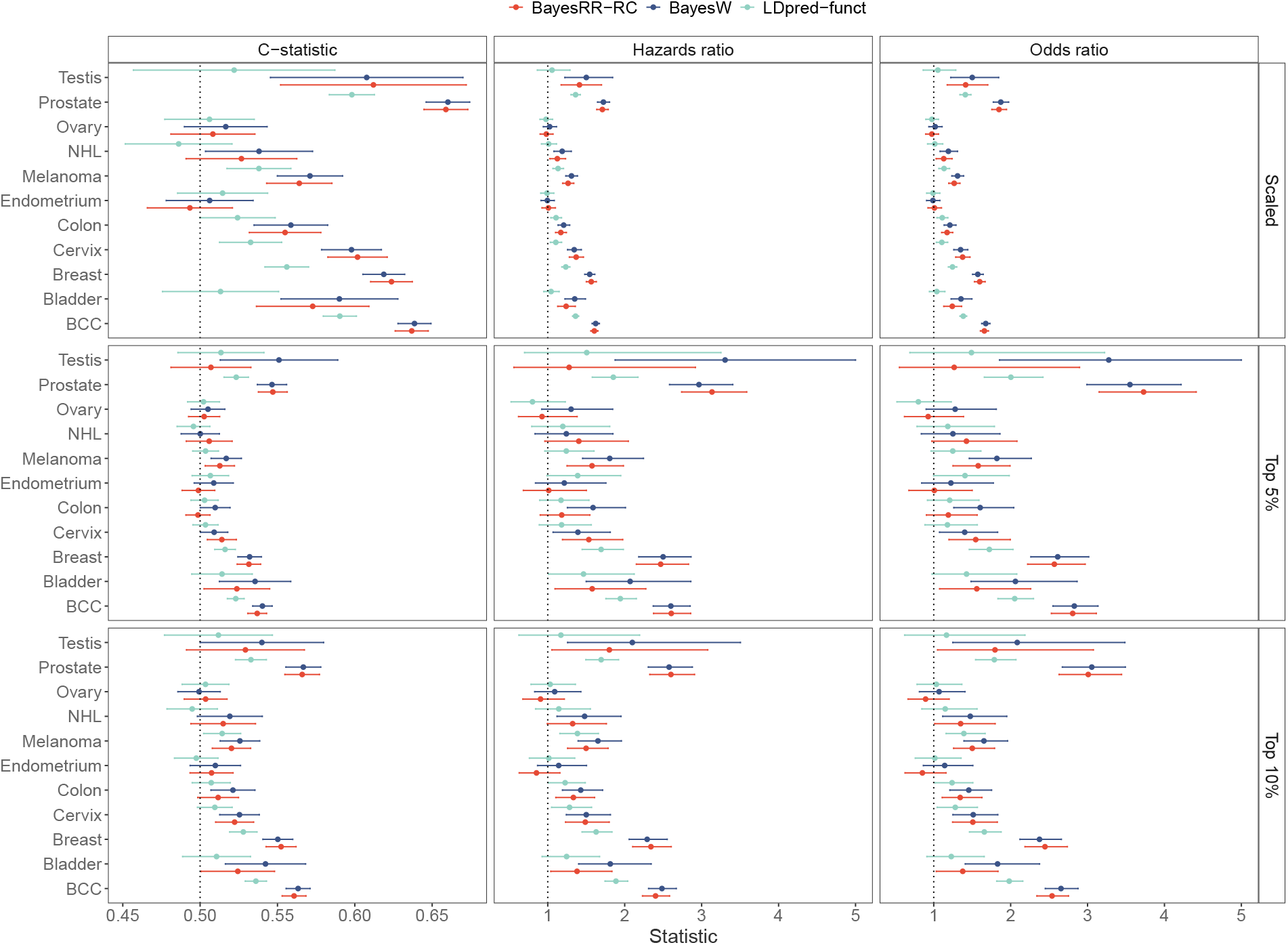
Predictive validation of different PRS on Estonian Biobank data using Harrell’s C-statistic, hazards ratio or odds ratio with 95% CI. The statistics were calculated by finding the impact of one standard deviation increase in the PRS (Scaled), by finding the impact of belonging to top 5% quantile of the PRS or by finding the impact of belonging to the top 10% quantile of the PRS on the likelihood of having cancer. Harrel’s C-statistic was calculated from Cox proportional hazards model without covariates, odds ratio was calculated from a logistic model using sex and age-at-entry as covariates, hazards ratio was calculated from Cox proportional hazards model using sex and age-at-entry as covariates.

**Figure S4.**
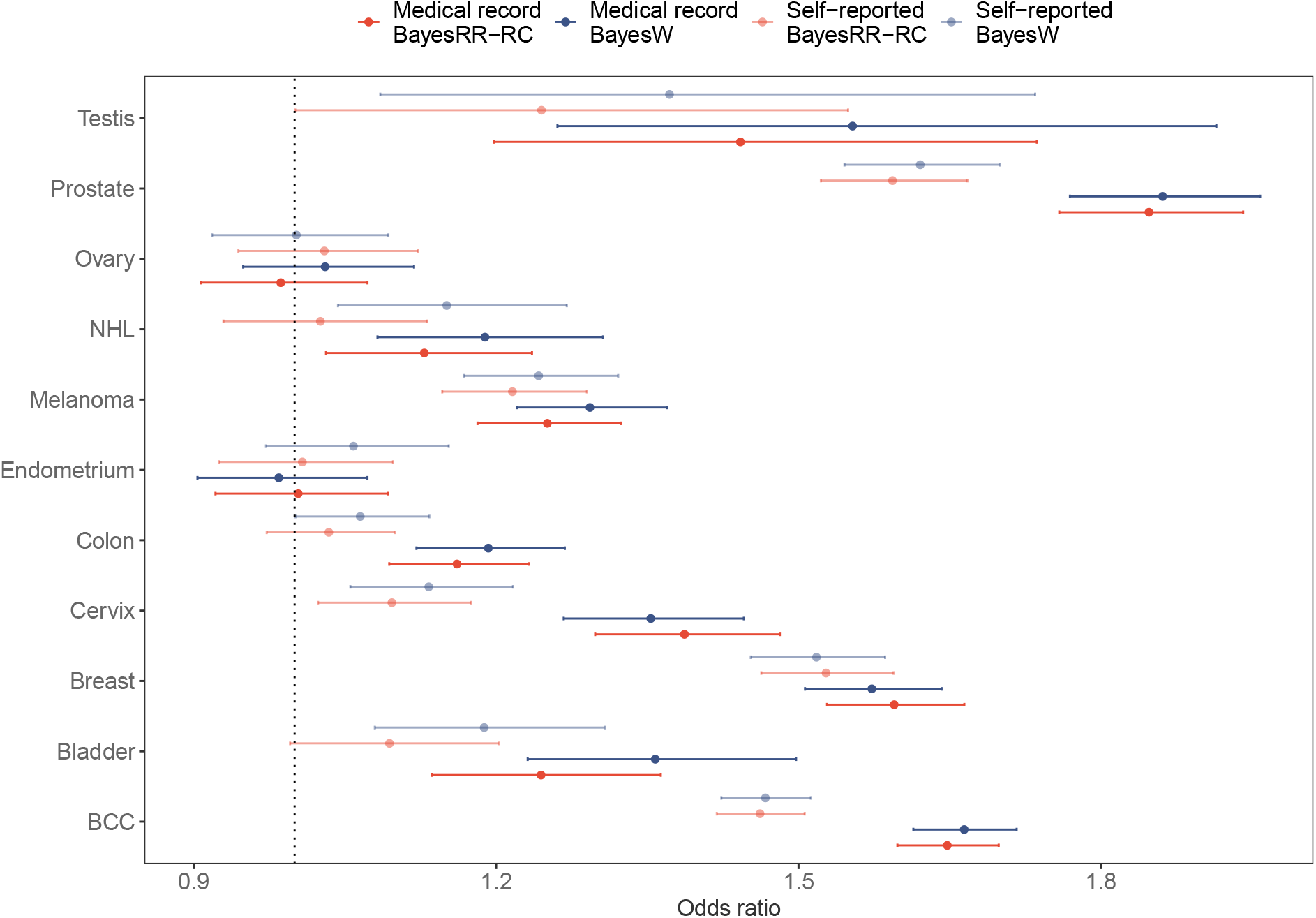
Prediction in Estonian Biobank using either medical record or self-reported phenotypic data in BayesW or BayesRR-RC models. The polygenic risk scores that are using medical record data rather than self-reported data tend to be more predictive across all cancers. The odds ratios were calculated by finding the impact of one standard deviation increase in PRS in a logistic model using sex and age-at-entry as covariates.

**Figure S5.**
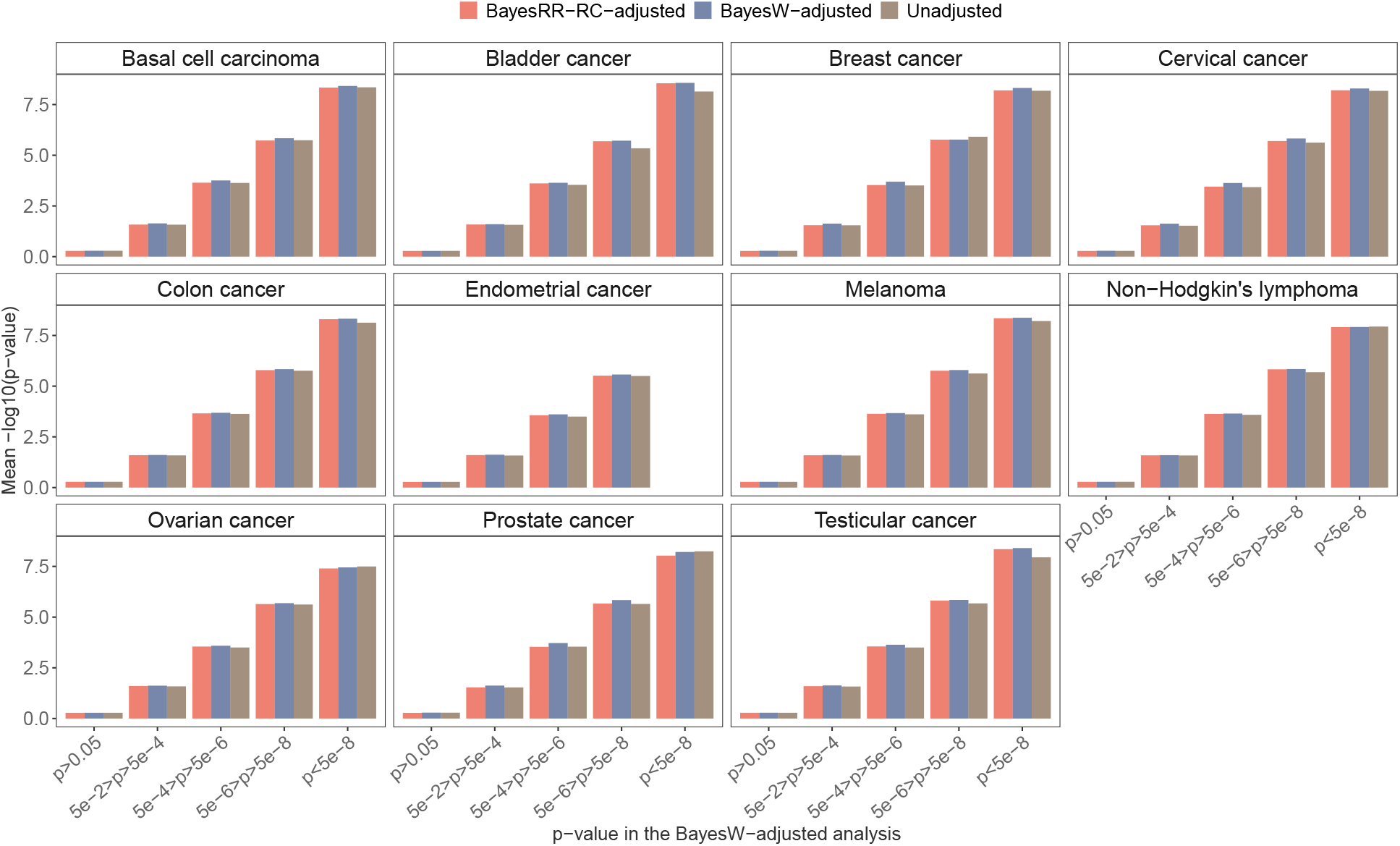
Mean -log10 p-value from the marginal association analysis adjusted with either BayesRR-RC, BayesW or without adjustment. The significance of each SNP was obtained using a logistic regression score test from step 2 of REGENIE on binary (case-control) phenotypes. We observe that BayesW or BayesRR-RC LOCO adjustments result in similar or decreased p-values suggesting increased statistical power.

**Figure S6.**
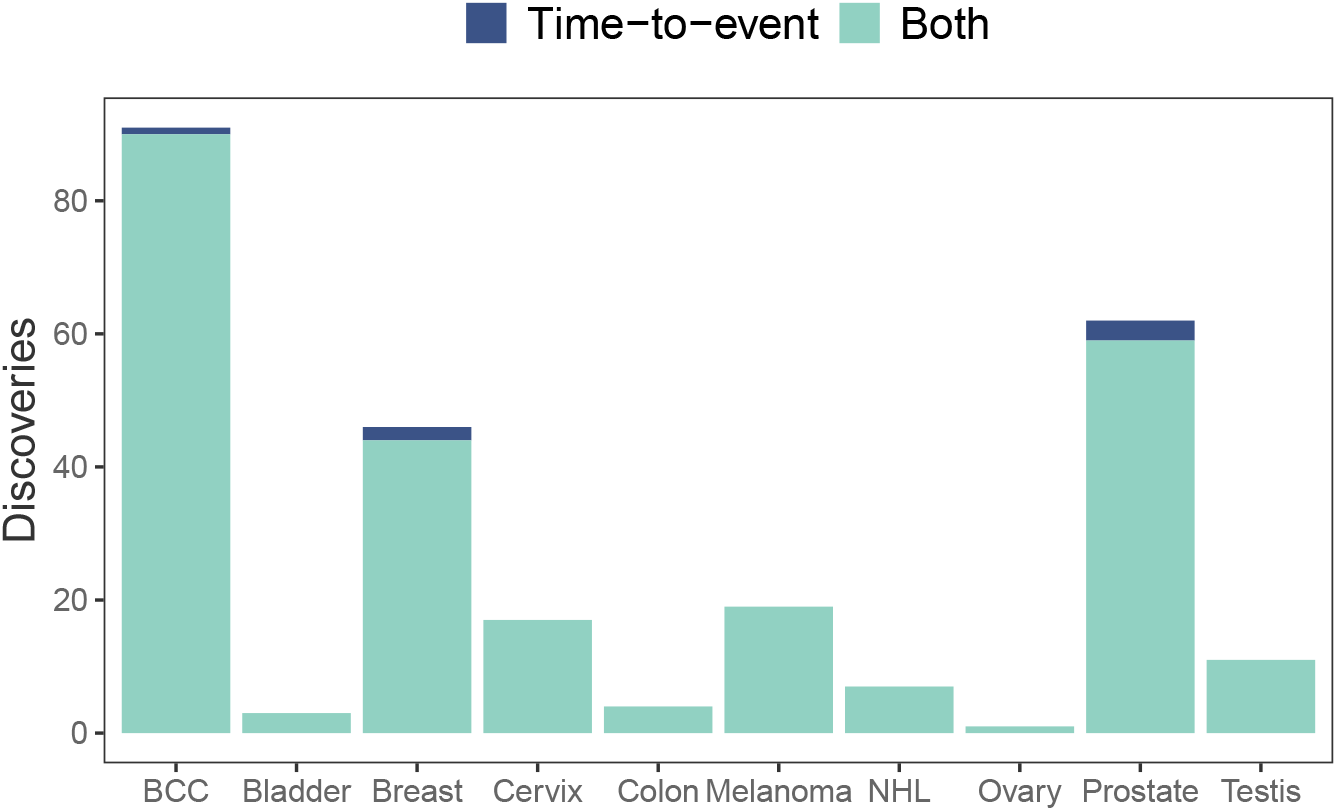
Classification of previously reported discoveries by each cancer type. Case-control and time-to-event substantially overlap in recovering previous findings (255/261). However, time-to-event adjustment enables replicating 6 additional loci, 1 for basal cell carcinoma, 2 for breast cancer and 3 for prostate cancer. All case-control approach discoveries were replicated by the time-to-event approach discovery.

**Figure S7.**
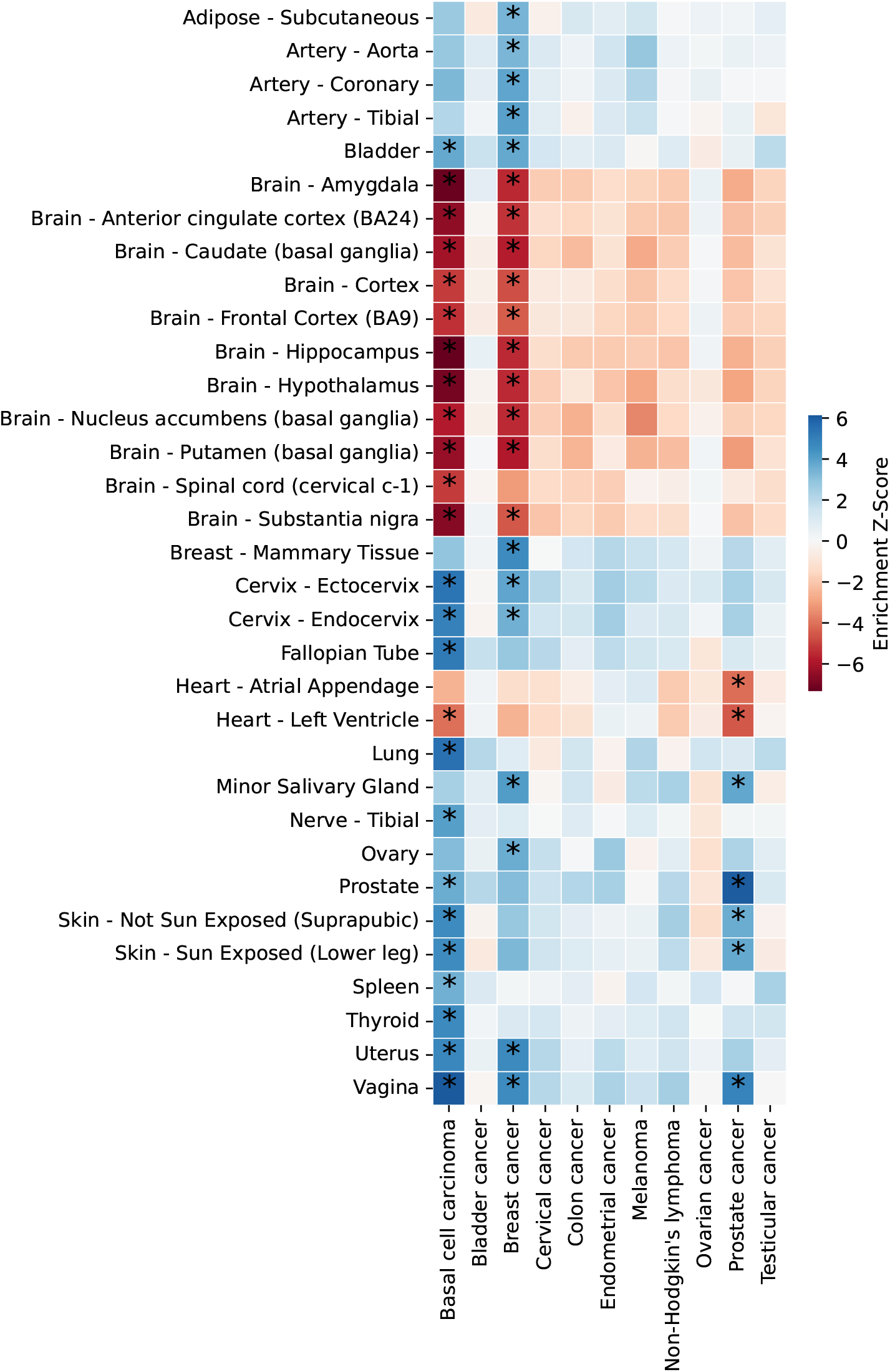
Tissue-specific enrichment for GTEx v8 tissues. For basal cell carcinoma, breast and prostate cancer, Downstreamer analysis highlighted significant enrichment in several tissues, including tissue-specific associations: e.g. in both sun-exposed and not sun-exposed skin for basal cell carcinoma, in mammary tissue for breast cancer, and prostate for prostate cancer. Enrichment Z-scores that were both Bonferroni and 5% FDR significant are marked with asterisks. Only tissues with significant enrichment Z-scores are shown.

